# Facilitating safe discharge through predicting disease progression in moderate COVID-19: a prospective cohort study to develop and validate a clinical prediction model in resource-limited settings

**DOI:** 10.1101/2021.12.02.21267170

**Authors:** Arjun Chandna, Raman Mahajan, Priyanka Gautam, Lazaro Mwandigha, Karthik Gunasekaran, Divendu Bhusan, Arthur T L Cheung, Nicholas Day, Sabine Dittrich, Arjen Dondorp, Tulasi Geevar, Srinivasa R Ghattamaneni, Samreen Hussain, Carolina Jimenez, Rohini Karthikeyan, Sanjeev Kumar, Shiril Kumar, Vikash Kumar, Debasree Kundu, Ankita Lakshmanan, Abi Manesh, Chonticha Menggred, Mahesh Moorthy, Jennifer Osborn, Melissa Richard-Greenblatt, Sadhana Sharma, Veena K Singh, Vikash K Singh, Javvad Suri, Shuichi Suzuki, Jaruwan Tubprasert, Paul Turner, Annavi M G Villanueva, Naomi Waithira, Pragya Kumar, George M Varghese, Constantinos Koshiaris, Yoel Lubell, Sakib Burza

**Author notes:** **CORRESPONDING AUTHORS** Dr. Arjun Chandna, Dr. Sakib Burza.

## Abstract

**Background:** In locations where few people have received COVID-19 vaccines, health systems remain vulnerable to surges in SARS-CoV-2 infections. Tools to identify patients suitable for community-based management are urgently needed.

**Methods:** We prospectively recruited adults presenting to two hospitals in India with moderate symptoms of laboratory-confirmed COVID-19 in order to develop and validate a clinical prediction model to rule-out progression to supplemental oxygen requirement. The primary outcome was defined as any of the following: SpO_2_ < 94%; respiratory rate > 30 bpm; SpO_2_/FiO_2_ < 400; or death. We specified *a priori* that each model would contain three clinical parameters (age, sex and SpO_2_) and one of seven shortlisted biochemical biomarkers measurable using near-patient tests (CRP, D-dimer, IL-6, NLR, PCT, sTREM-1 or suPAR), to ensure the models would be suitable for resource-limited settings. We evaluated discrimination, calibration and clinical utility of the models in a temporal external validation cohort.

**Findings:** 426 participants were recruited, of whom 89 (21·0%) met the primary outcome. 257 participants comprised the development cohort and 166 comprised the validation cohort. The three models containing NLR, suPAR or IL-6 demonstrated promising discrimination (c-statistics: 0·72 to 0·74) and calibration (calibration slopes: 1·01 to 1·05) in the validation cohort, and provided greater utility than a model containing the clinical parameters alone.

**Interpretation:** We present three clinical prediction models that could help clinicians identify patients with moderate COVID-19 suitable for community-based management. The models are readily implementable and of particular relevance for locations with limited resources.

**Funding:** Médecins Sans Frontières, India.

**RESEARCH IN CONTEXT:** *Evidence before this study:* A living systematic review by Wynants et al. identified 137 COVID-19 prediction models, 47 of which were derived to predict whether patients with COVID-19 will have an adverse outcome. Most lacked external validation, relied on retrospective data, did not focus on patients with moderate disease, were at high risk of bias, and were not practical for use in resource-limited settings. To identify promising biochemical biomarkers which may have been evaluated independently of a prediction model and therefore not captured by this review, we searched PubMed on 1 June 2020 using synonyms of “SARS-CoV-2” AND [“biomarker” OR “prognosis”]. We identified 1,214 studies evaluating biochemical biomarkers of potential value in the prognostication of COVID-19 illness. In consultation with FIND (Geneva, Switzerland) we shortlisted seven candidates for evaluation in this study, all of which are measurable using near-patient tests which are either currently available or in late-stage development.

*Added value of this study:* We followed the TRIPOD guidelines to develop and validate three promising clinical prediction models to help clinicians identify which patients presenting with moderate COVID-19 can be safely managed in the community. Each model contains three easily ascertained clinical parameters (age, sex, and SpO_2_) and one biochemical biomarker (NLR, suPAR or IL-6), and would be practical for implementation in high-patient-throughput low resource settings. The models showed promising discrimination and calibration in the validation cohort. The inclusion of a biomarker test improved prognostication compared to a model containing the clinical parameters alone, and extended the range of contexts in which such a tool might provide utility to include situations when bed pressures are less critical, for example at earlier points in a COVID-19 surge.

*Implications of all the available evidence:* Prognostic models should be developed for clearly-defined clinical use-cases. We report the development and temporal validation of three clinical prediction models to rule-out progression to supplemental oxygen requirement amongst patients presenting with moderate COVID-19. The models are readily implementable and should prove useful in triage and resource allocation. We provide our full models to enable independent validation.

## INTRODUCTION

As COVID-19 continues to spread, global attention is moving towards the safe ‘re-opening’ of societies and borders. In low-income countries, where fewer than 5% of people have received a COVID-19 vaccine (Figure 1),^1^ fragile healthcare systems remain vulnerable to being overwhelmed by a surge in COVID-19 cases.^2-4^

**Figure 1.**
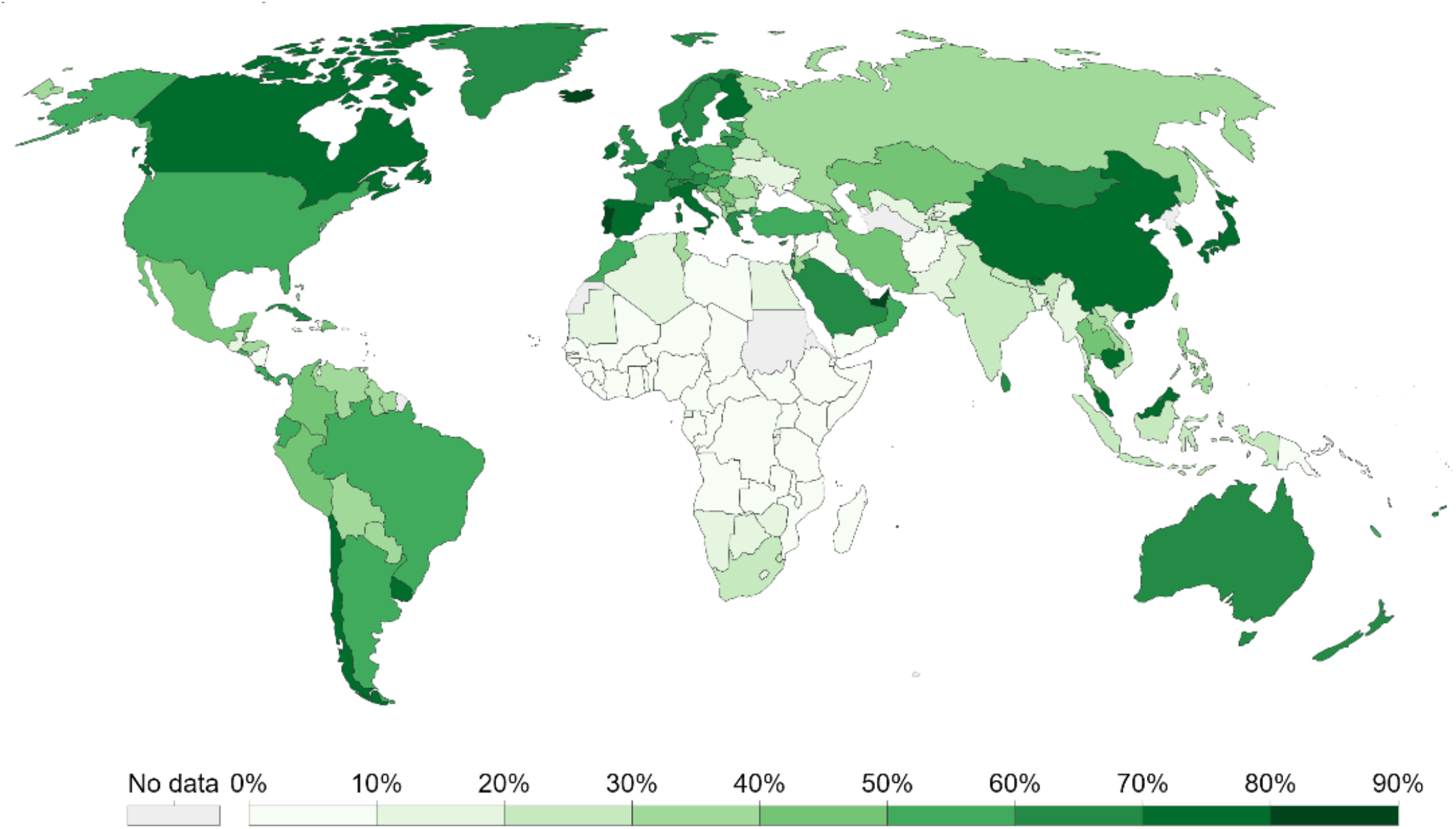
Proportion of individuals fully vaccinated against COVID-19 as of 7 November 2021. Adapted from ourworldindata.org.^1^

Only a small proportion of patients with COVID-19 require admission to hospital. Oxygen is the most important supportive treatment and in most low- and middle-income countries (LMICs) is the practical ceiling of care.^5^ The World Health Organization (WHO) estimates that 15% of patients with symptomatic COVID-19 will require supplemental oxygen.^6^ Effective identification of patients who are unlikely to become hypoxic and suitable for community-based management would have considerable benefit; tools to support triage could decompress healthcare systems by giving practitioners confidence to allocate resources more efficiently.^7^ Efficient and accurate allocation becomes increasingly important as resources are stretched by high case numbers.

Numerous prognostic models for COVID-19 have been developed.^8,9^ Almost all predict critical illness or mortality and thus cannot inform whether a patient might be safely managed in the community. Of the few that focus on patients with moderate disease, most rely on retrospective or registry-based data,^10-14^ lack external validation,^15,16^ and are not feasible for use in resource-limited settings.^9,17^ Moreover, most existing studies did not follow best-practice guidelines for model building and reporting,^18^ are at high risk of bias,^8^ and the resulting models are neither suitable nor recommended for use in LMIC contexts.^9^

We set out to develop and validate a clinical prediction model to rule-out progression to supplemental oxygen requirement in patients presenting with moderate COVID-19 (PRIORITISE; Prognostication of Oxygen Requirement in Patients with Non-severe SARS-CoV-2 Infection). We hypothesised that combining simple clinical parameters with host biomarkers feasible for measurement in resource-limited settings and implicated in the pathogenesis of COVID-19 would improve prognostication. The study was motivated by a recognition that a tool to inform this critical clinical decision could support healthcare systems in under-resourced settings during current and future surges in SARS-CoV-2 infections.^2^

## METHODS

### Study population

PRIORITISE is a prospective observational cohort study. Consecutive patients aged ≥ 18 years with clinically-suspected SARS-CoV-2 infection presenting with moderate symptoms to the All India Institute of Medical Sciences (AIIMS) Hospital in Patna, India and the Christian Medical College (CMC) Hospital in Vellore, India were screened (daytime hours, Monday to Saturday). AIIMS is a 1,000-bed hospital and the largest medical facility providing primary-to-tertiary healthcare in the state of Bihar. CMC is a 3,000-bed not-for-profit medical college hospital that provided care for ∼1,500 patients with COVID-19 each day during the peak of the pandemic.

We adapted the case definitions in the WHO Clinical Management guideline (moderate disease)^6^ and WHO Clinical Progression Scale (WHO-CPS; scores 2, 3 or 4)^19^ to define moderate disease as follows: a peripheral oxygen saturation (SpO_2_) ≥ 94% and respiratory rate < 30 breaths per minute (bpm), in the context of systemic symptoms (breathlessness or fever and chest pain, abdominal pain, diarrhoea or severe myalgia), recognising that the threshold for hospitalisation varies throughout a pandemic and that a sensitive cut-off for hypoxia would be desirable in a tool to inform community-based management.^19,20^

### Data collection

Structured case-report forms were completed at enrolment, day 7, and day 14, and daily during admission for participants admitted to the study facilities. Anthropometrics and vital signs were measured at enrolment and demographics, clinical symptoms, comorbidities, and medication history collected via brief interview with the participant. Venous blood samples were collected at enrolment in ethylenediaminetetraacetic acid (EDTA) tubes. Participants were followed-up in-person whilst admitted to the facility, and by telephone on days 7 and 14 if discharged prior to this. Those discharged who reported worsening symptoms on day 7 and/or persistent symptoms on day 14 were recalled to have their SpO_2_ and respiratory rate measured.

### Primary outcome

The primary outcome was development of an oxygen requirement within 14 days of enrolment, defined as any of the following: SpO_2_ < 94%; respiratory rate > 30 bpm; SpO_2_/FiO_2_ < 400;^21,22^ or death, aligning closely with a WHO-CPS score of ≥ 5.^19^ Patients who received supplemental oxygen outside the study facilities were classified as meeting the primary outcome if it was not possible to retrieve their case notes, provided the oxygen was prescribed in a licensed medical facility. The site study teams were unaware of which baseline variables had been preselected as candidate predictors when determining outcome status.

### Candidate predictors

We decided *a priori* that a model using four predictors would be practical for use in high-patient-throughput resource-limited settings. Following review of the evidence and considering resource constraints, reliability, validity, feasibility (ease of collection, time-to-decision, etc.) and biological plausibility we prespecified that each model would contain age, sex, SpO_2_ and one biochemical biomarker.^10,17,23^

Following an initial scoping review (appendix p2-3), biomarkers were shortlisted in consultation with FIND, the global alliance for diagnostics (Geneva, Switzerland). To qualify for inclusion, biomarkers had to be quantifiable with near-patient tests in clinical use or late-stage development (Technology Readiness Level ≥ 4; appendix p4).^24^ The final list included seven biochemical biomarkers: C-reactive protein (CRP), D-dimer, interleukin-6 (IL-6), neutrophil-to-lymphocyte ratio (NLR), procalcitonin (PCT), soluble triggering receptor expressed on myeloid cells 1 (sTREM-1), and soluble urokinase plasminogen activator receptor (suPAR).^25-29^

Clinical predictors were measured at enrolment and all biomarkers except NLR were measured retrospectively from samples obtained at enrolment. NLR was measured on site and was not repeated if it had been measured at the site within 24 hours prior to recruitment. All predictors were measured blinded to outcome status.

### Laboratory procedures

Complete blood counts (XP-300-Hematology-Analyzer, Sysmex, IL) were performed on site and aliquots of EDTA-plasma stored at -20°C or below until testing. Plasma concentrations of biomarkers of immune activation, inflammation and coagulation were quantified using the suPARnostic ELISA (ViroGates, Lyngby) and Simple Plex Ella microfluidic platform (ProteinSimple, CA) as described elsewhere.^30^ Remaining plasma was biobanked on site. SARS-CoV-2 IgG and IgM antibodies were measured using the SCoV-2 Detect ELISA (InBios, WA). Combined oral and/or nasopharyngeal swabs were collected to confirm SARS-CoV-2 infection via RT-PCR (Cepheid Xpert Xpress SARS-CoV-2, CA or Altona RealStar SARS-CoV-2 rRT-PCR, Hamburg).

### Sample size

We followed the recommendations of Riley et al. and assumed a conservative R^2^ Nagelkerke of 0·15.^31^ We anticipated that ∼8% of participants would meet the primary endpoint and estimated that 44 outcome events would be required to derive a prediction model comprising four candidate predictors (events per parameter [EPP] = 11) and minimise the risk of overfitting. Allowing for 5% attrition, we aimed to recruit 600 participants in the model development cohort.

Given the uncertainty around deterioration rates amongst patients with moderate COVID-19 at the time of study inception, we prespecified an interim review after the first 100 participants were recruited. At this review, the proportion of participants meeting the primary endpoint was higher than anticipated (20% vs. 8%). At this higher prevalence, and using R^2^ values from 0·20-0·15, between 52-68 outcome events (EPP = 13-17) would be required to develop the prediction models.^31^ Recognising that (i) our range of R^2^ estimates was conservative, (ii) penalised regression methods would reduce the risk of overfitting, and (iii) the external validation cohort would allow assessment of model optimism, and following the advice of the Study Management Group and External Advisory Panel, a decision was made to use the first 50 outcome events to derive the models. Participants recruited after that point were entered into the external temporal validation cohort.

### Model development and validation

We used logistic regression to develop the models given the short time horizon (14-day). Ridge regression was used to shrink regression coefficients and minimise model optimism. All predictors were prespecified and no predictor selection was performed during model development. The relationship between each continuous predictor (age, SpO_2_, all biomarkers) and the primary outcome was examined and transformations used as appropriate. Due to the small fraction of missing data (< 3% for any single predictor), missing observations were replaced with the median value, grouped by outcome status. A sensitivity analysis was conducted using full-case analysis.

We assessed discrimination (c-statistic; how well participants who met the primary outcome are differentiated from those who did not) and calibration (calibration plots and slopes; agreement between predicted probabilities and observed outcomes) for each model in the validation cohort.

Next, given that our aim was to develop a model to rule-out progression to oxygen requirement, we examined classifications (true positives [TP], false positives [FP], true negatives [TN], false negatives [FN]) at clinically-relevant cut-points (predicted probabilities). Finally, recognising that the relative value of a TP and FP will vary at different stages of the pandemic (for example, reflecting bed pressures and/or capacity for follow-up),^20^ we examined the potential clinical utility of the models using decision curve analyses to quantify the net benefit between correctly identified TP or TN and incorrectly identified FP or FN at a range of plausible trade-offs (threshold probabilities).^32^ We were particularly interested in the threshold probabilities at which the prediction models might offer benefit over the “admit-all” approach and the added value of the models containing each of the candidate biomarkers compared to the clinical model.

All analyses were done in R v4.03.

### Ethical approvals

This investigator-initiated study was prospectively registered (ClinicalTrials.gov; NCT04441372), with protocol and statistical analysis plan uploaded to the Open Science Framework platform (DOI: 10.17605/OSF.IO/DXQ43). Ethical approval was given by the AIIMS, Patna Ethics Committee; CMC Ethics Committee; Oxford Tropical Research Ethics Committee; and MSF Ethical Review Board.

## RESULTS

Between 22 October 2020 and 3 July 2021, 2,808 patients with clinically-suspected COVID-19 were screened, of whom 446 were eligible (446/2,808; 15·9%) and 426 were recruited (20/446; 4·5% refusal rate). Three participants were lost-to-follow-up (3/426; 0·7%) and excluded from further analyses (Figure 2). All participants had laboratory-confirmed SARS-CoV-2 infection (421/423 [99·5%] via RT-PCR). The maximum amount of missing data for any predictor was 2·6% (NLR; 11/423; appendix p5). The first 257 participants comprised the development cohort and the remaining 166 participants comprised the temporal validation cohort.

**Figure 2.**
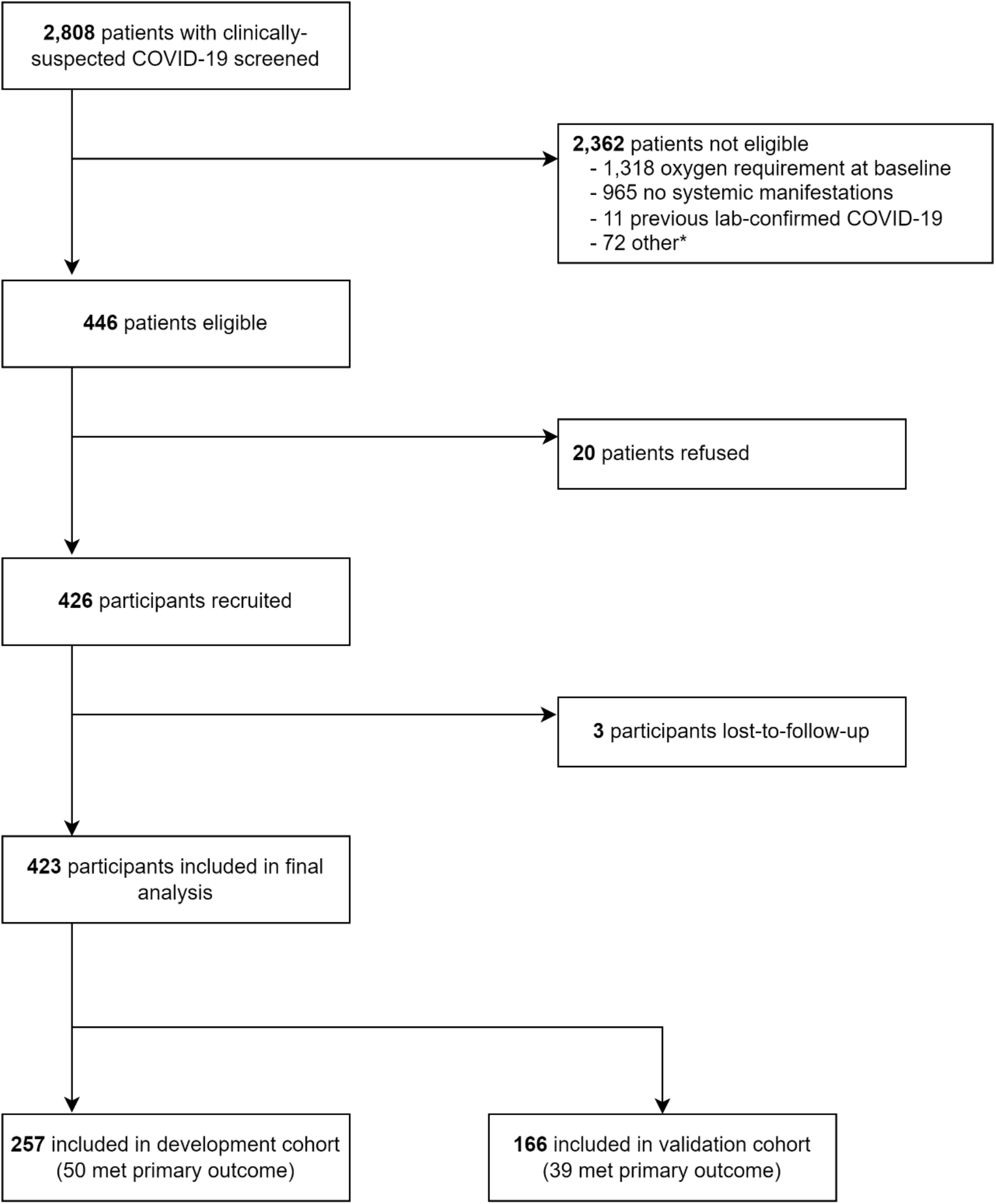
Screening and recruitment of participants into the PRIORITSE study. ^*^Reasons for exclusion: 64 vaccinated, 3 unable to provide consent, and 5 reason not documented. Towards the end of recruitment (March 2021 in AIIMS and May 2021 in CMC) vaccines against COVID-19 began to be rolled out in the study areas and a decision was made to exclude vaccinated participants as the study would not be powered to determine whether the prediction models were valid in this cohort.

### Outcomes

Development and validation cohorts were largely balanced with respect to baseline characteristics (Table 1; appendix p6-9). There was a higher proportion of males in the development cohort (72% [185/257] vs. 61% [101/166]). In the validation cohort, more participants had a qSOFA score ≥ 2 (16/166 [9·6%] vs. 13/257 [5·1%]), and the validation cohort had higher median CRP (58·1 mg/l vs. 24·4 mg/l) and IL-6 (31·6 pg/ml vs. 11·0 pg/ml) concentrations. Eighty-nine participants met the primary outcome (89/423; 21·0%); 50 in the development cohort (50/257; 19·5%) and 39 in the validation cohort (39/166; 23·5%). The median (IQR) time to oxygen requirement was 1 (1-3) day; 11 participants died, 2 were mechanically ventilated, 15 received non-invasive ventilation, 49 received oxygen via a face mask and/or nasal cannula (one outside the study facilities), and 12 had an SpO_2_ < 94% but did not receive oxygen supplementation (appendix p10-11).

**Table 1.**
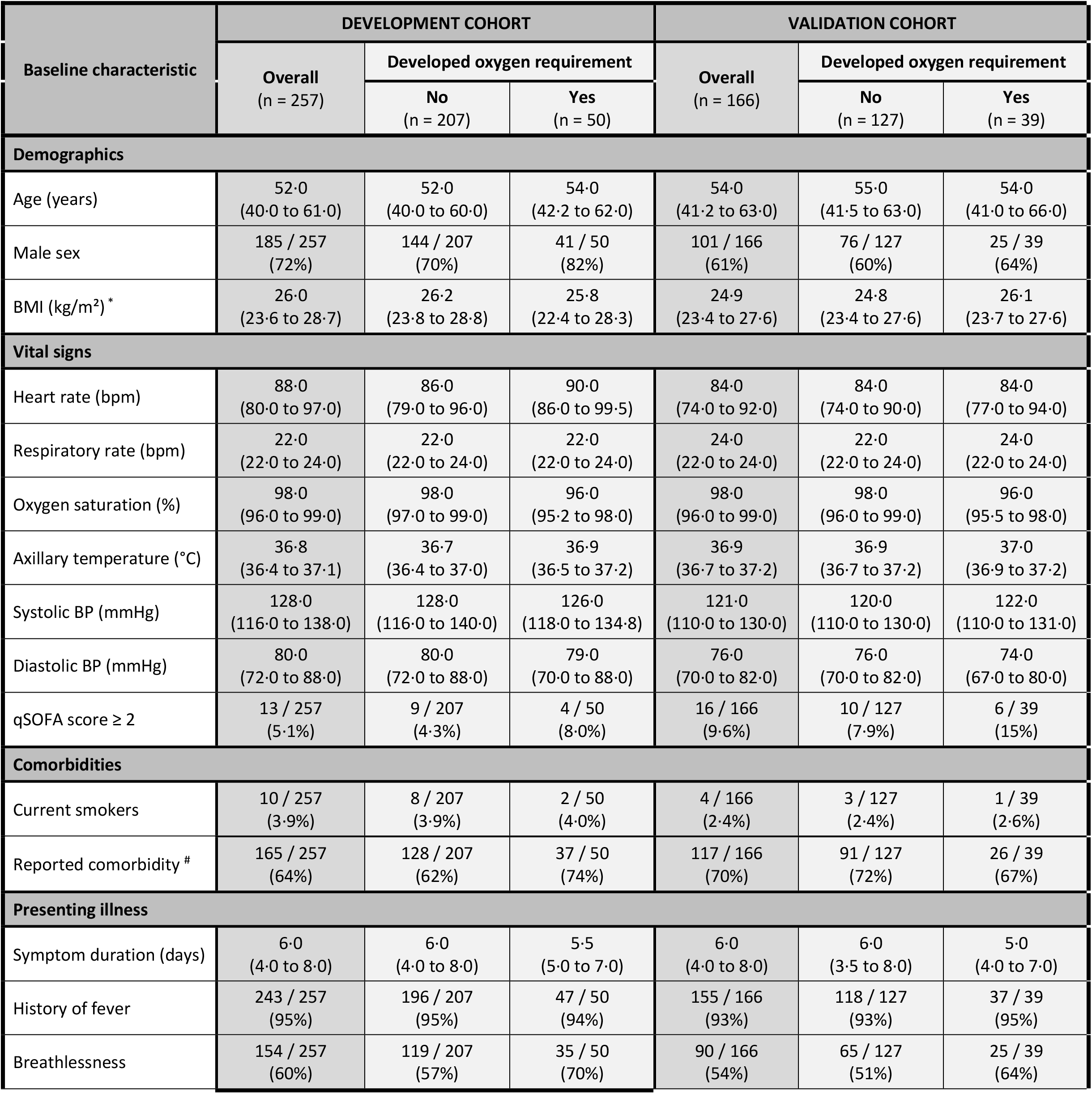
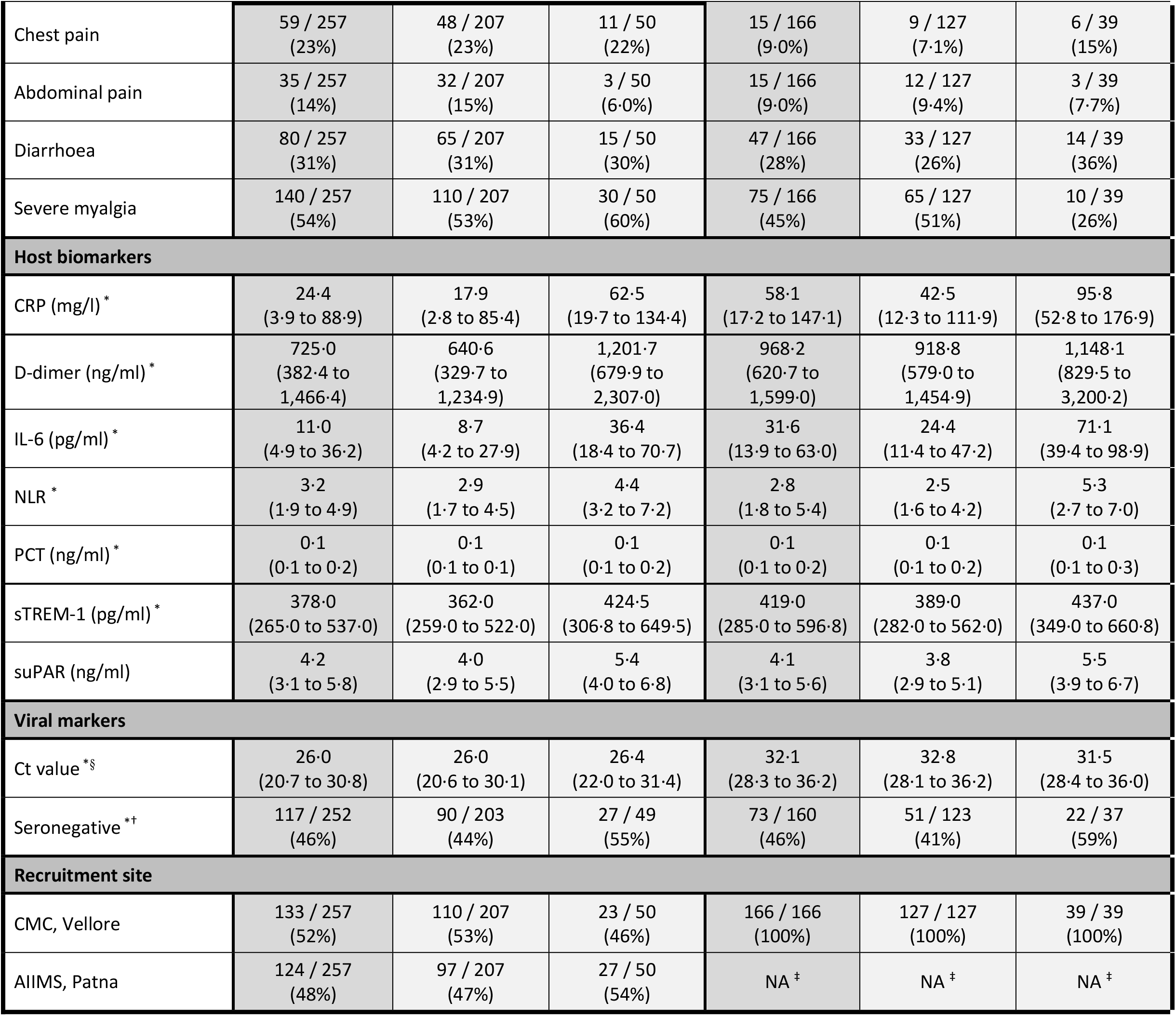
Baseline characteristics of development and validation cohorts, stratified by primary outcome status. ^*^Missing data: BMI = 1; CRP = 8, D-dimer = 3, IL-6 = 2, NLR = 12; PCT = 2; sTREM-1 = 2; Ct value = 181; serostatus = 11. ^#^Details of the 12 comorbidities that participants were asked about are provided in appendix p6-9. Comorbidities are not reliably diagnosed or known by patients in our contexts and therefore were not selected as one of the *a priori* clinical predictors. ^§^Different specimen collection procedures and PCR assays were used at each site (appendix p21). ^†^Seronegative defined as negative for both SARS-CoV-2 IgG and IgM antibodies. ^‡^Recruitment closed in Patna in March 2021 and only participants from CMC were recruited into the temporal validation cohort. Median values (IQR) are reported for continuous variables.

Associations between candidate predictors and the primary outcome are illustrated (Figure 3), and c-statistics (continuous predictors) and odds ratios (continuous and categorical predictors) reported (appendix p12-14), although univariate associations were not used to inform predictor selection for the modelling. The full models, including intercepts, regression coefficients and adjusted odds ratios, are presented in the appendix (p12-14). After adjustment for the three clinical variables, five biomarkers (CRP, D-dimer, IL-6, NLR, and suPAR) were independently associated with the development of an oxygen requirement.

**Figure 3.**
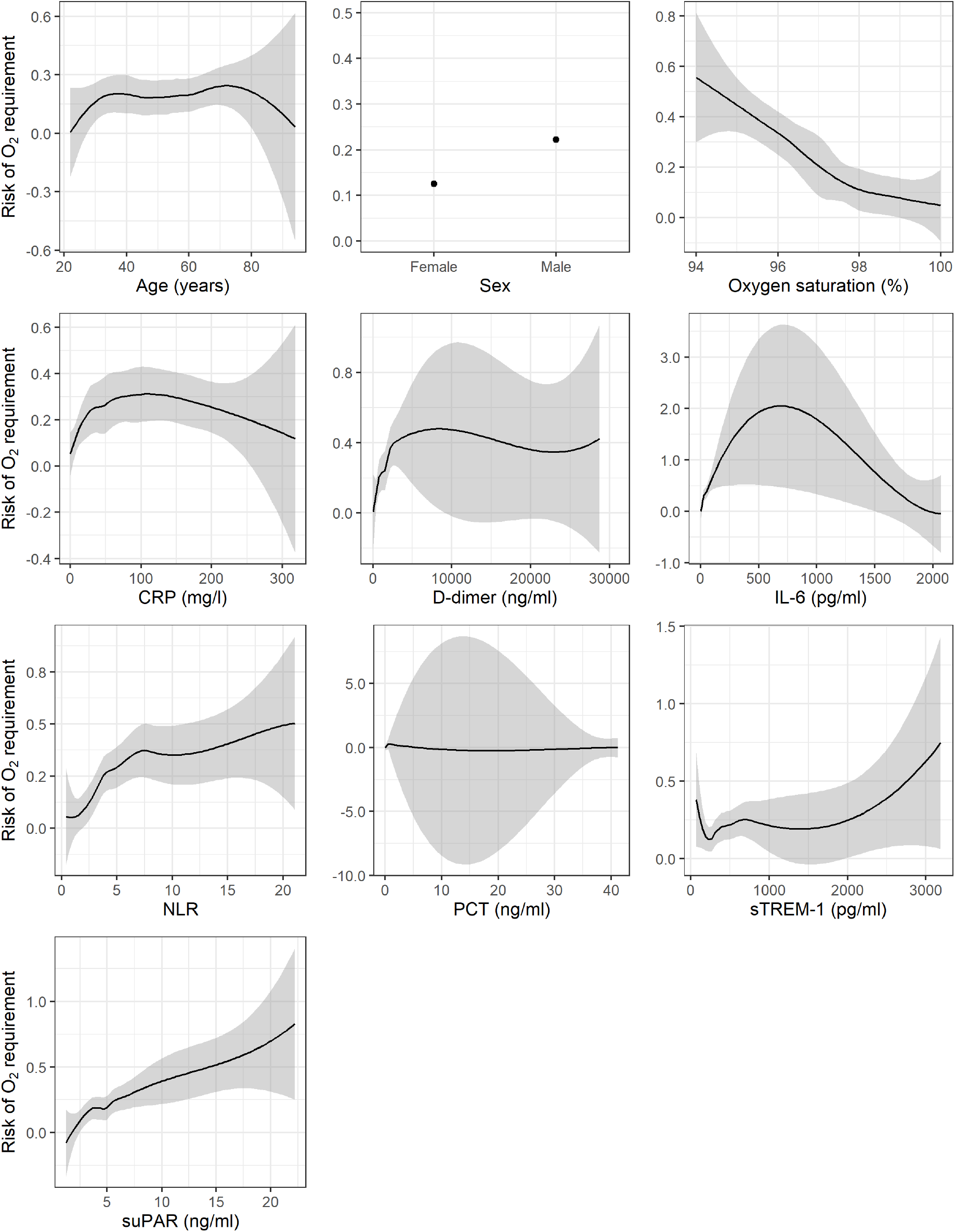
Univariable associations between candidate predictors and primary outcome in the development cohort. Dots and lines indicate point estimates; grey shaded areas indicate 95% confidence intervals.

### Prognostic models

Discrimination and calibration of each model in the temporal validation cohort are presented in Figure 4. C-statistics ranged from 0·66 (clinical model and model containing PCT) to 0·74 (model containing IL-6). Calibration slopes ranged from 0·62 (model containing PCT) to 1·01 (model containing suPAR). Calibration was better at lower predicted probabilities, with some models overestimating risk at higher predicted probabilities.

**Figure 4.**
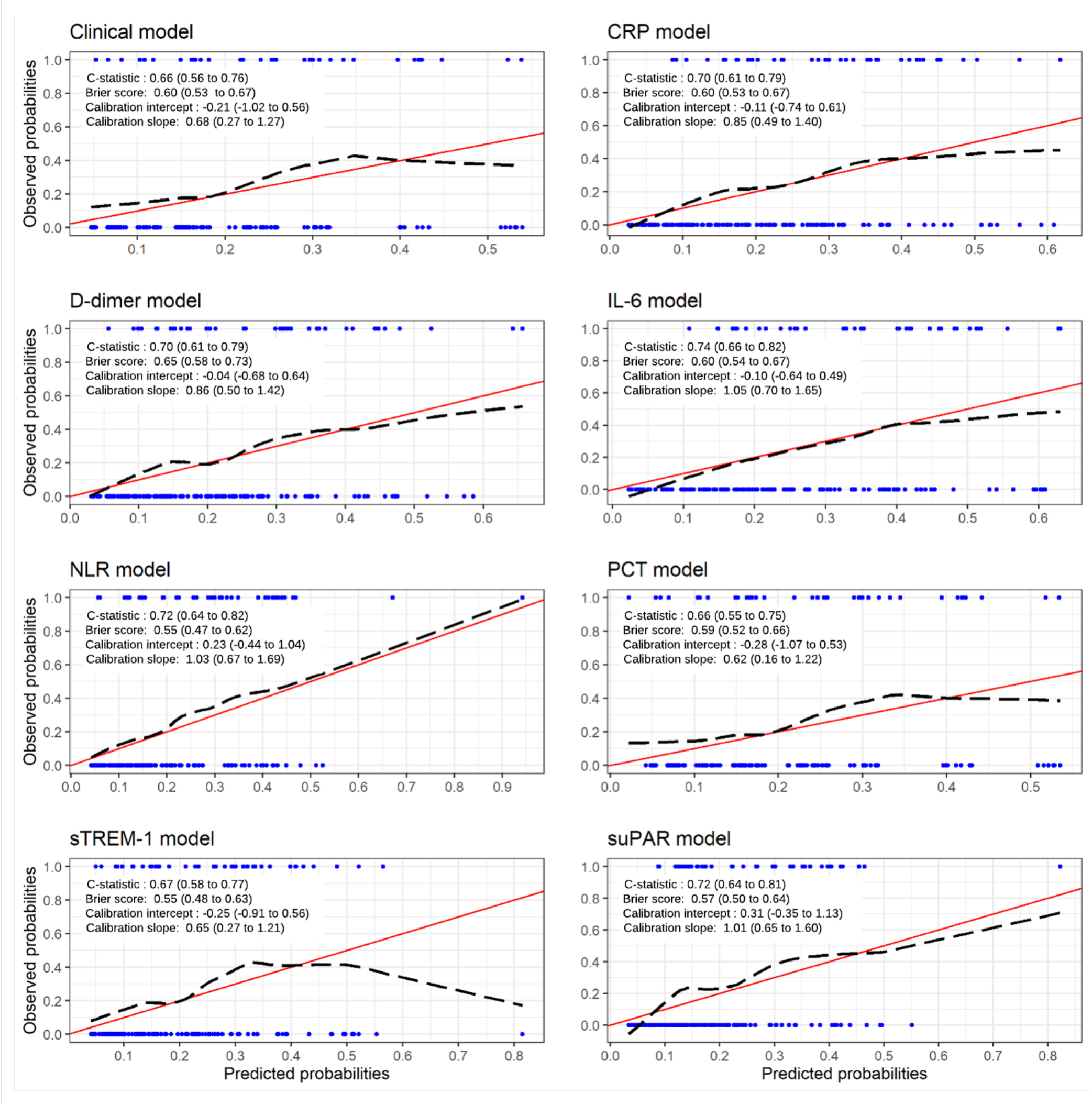
Performance measures and calibration plots for each model in the validation cohort. Red line indicates perfect calibration; black dashed line indicates calibration slope for that particular model; blue rug plots indicate distribution of predicted risk for participants who did (top) and did not (bottom) meet the primary outcome. C-statistics indicate how well participants who met the primary outcome are differentiated from those who did not; perfect discrimination is indicated by a c-statistic of 1·0. Calibration slopes indicate agreement between predicted probabilities and observed outcomes; perfect calibration is indicated by a slope of 1·0.

The ability of each model to rule-out progression to oxygen requirement amongst patients with moderate COVID-19 at predicted probabilities (cut-offs) of 10%, 15% and 20% is shown (Table 2; appendix p15-18). A cut-off of 10% reflects a management strategy equivalent to admitting any patient in whom the predicted risk of developing an oxygen requirement is ≥ 10%. At this cut-off, the results suggest that a model containing the three clinical parameters (age, sex, and SpO_2_) without any biomarkers could facilitate correctly sending home ∼25% of patients with moderate COVID-19 who would not subsequently require supplemental oxygen, at the cost of also sending home ∼9% of moderate patients who would deteriorate and require supplemental oxygen, i.e. a ratio of correctly to incorrectly discharged patients of 10:1.

**Table 2.** Predicted classification of patients at different cut-offs for each model, using the prevalence of the primary outcome in the validation cohort. A cut-off of 0·1 reflects a management strategy in which any patient with a predicted risk of requiring oxygen ≥ 10% is admitted. FN = false negative; FP = false positive; TN = true negative; TP = true positive.

| Predicted probability of model (cut-off) | Per 100 patients (23 patients who would require oxygen) |  |  |  | Ratio of incorrect to correct admissions (FP : TP) | Ratio of correct to incorrect discharges (TN : FN) |
| --- | --- | --- | --- | --- | --- | --- |
|  | Patients who would require oxygen admitted (TP) | Unnecessary hospital admissions (FP) | Patients who would require oxygen discharged (FN) | Patients correctly discharged (TN) |  |  |
| Clinical model |  |  |  |  |  |  |
| 0.1 | 21 | 58 | 2 | 19 | 3 to 1 | 10 to 1 |
| 0.15 | 18 | 46 | 5 | 31 | 3 to 1 | 6 to 1 |
| 0.20 | 14 | 29 | 9 | 48 | 2 to 1 | 5 to 1 |
| IL-6 model |  |  |  |  |  |  |
| 0.1 | 23 | 61 | 0 | 16 | 3 to 1 | NA |
| 0.15 | 21 | 49 | 2 | 28 | 2 to 1 | 14 to 1 |
| 0.20 | 19 | 38 | 4 | 39 | 2 to 1 | 10 to 1 |
| NLR model |  |  |  |  |  |  |
| 0.1 | 22 | 54 | 1 | 23 | 2 to 1 | 23 to 1 |
| 0.15 | 17 | 39 | 6 | 38 | 2 to 1 | 6 to 1 |
| 0.20 | 15 | 25 | 8 | 52 | 2 to 1 | 6 to 1 |
| suPAR model |  |  |  |  |  |  |
| 0.1 | 22 | 52 | 1 | 25 | 2 to 1 | 25 to 1 |
| 0.15 | 16 | 34 | 7 | 43 | 2 to 1 | 6 to 1 |
| 0.20 | 13 | 22 | 10 | 55 | 2 to 1 | 6 to 1 |
| CRP model |  |  |  |  |  |  |
| 0.1 | 21 | 54 | 2 | 23 | 3 to 1 | 12 to 1 |
| 0.15 | 20 | 43 | 3 | 34 | 2 to 1 | 11 to 1 |
| 0.20 | 16 | 36 | 7 | 41 | 2 to 1 | 6 to 1 |
| D-dimer model |  |  |  |  |  |  |
| 0.1 | 21 | 54 | 2 | 23 | 3 to 1 | 12 to 1 |
| 0.15 | 19 | 39 | 4 | 38 | 2 to 1 | 10 to 1 |
| 0.20 | 15 | 31 | 8 | 46 | 2 to 1 | 6 to 1 |
| PCT model |  |  |  |  |  |  |
| 0.1 | 21 | 57 | 2 | 20 | 3 to 1 | 10 to 1 |
| 0.15 | 18 | 45 | 5 | 32 | 2 to 1 | 6 to 1 |
| 0.20 | 14 | 27 | 9 | 50 | 2 to 1 | 6 to 1 |
| sTREM-1 model |  |  |  |  |  |  |
| 0.1 | 20 | 55 | 3 | 22 | 3 to 1 | 7 to 1 |
| 0.15 | 17 | 41 | 6 | 36 | 2 to 1 | 6 to 1 |
| 0.20 | 14 | 28 | 9 | 49 | 2 to 1 | 5 to 1 |

The inclusion of either NLR or suPAR improved the predictive performance such that the ratio of correctly to incorrectly discharged patients increased to 23:1 or 25:1 respectively, whilst a model containing IL-6 resulted in a similar proportion (∼21%) of correctly discharged patients as the clinical model but without missing any patients who would deteriorate and require supplemental oxygen. Inclusion of any of the other candidate biomarkers (CRP, D-dimer, PCT or sTREM-1) did not improve the ability of the clinical model to rule-out progression to supplemental oxygen requirement.

### Generalisability

We recognised that the relative value of a TP and FP, i.e. admitted patients who would and would not subsequently require supplemental oxygen, was not fixed and would likely vary at different stages of the pandemic, reflecting bed pressures and/or capacity for follow-up.^20^ Decision curve analyses accounting for this differential weighting suggest that the clinical model could provide utility (net benefit over an “admit-all” approach) at a threshold probability above 15% (i.e. when the value of one TP is equal to ∼7 FPs). Furthermore, the results indicate that models containing any one of IL-6, NLR or suPAR could offer greater net benefit than the clinical model and extend the range of contexts in which a model might provide utility to include threshold probabilities above 5% (value of one TP is equal to 19 FPs; i.e. when bed pressures are less critical). For the model containing IL-6, this higher net benefit appeared to be maintained across a range of plausible threshold probabilities (Figure 5).

**Figure 5.**
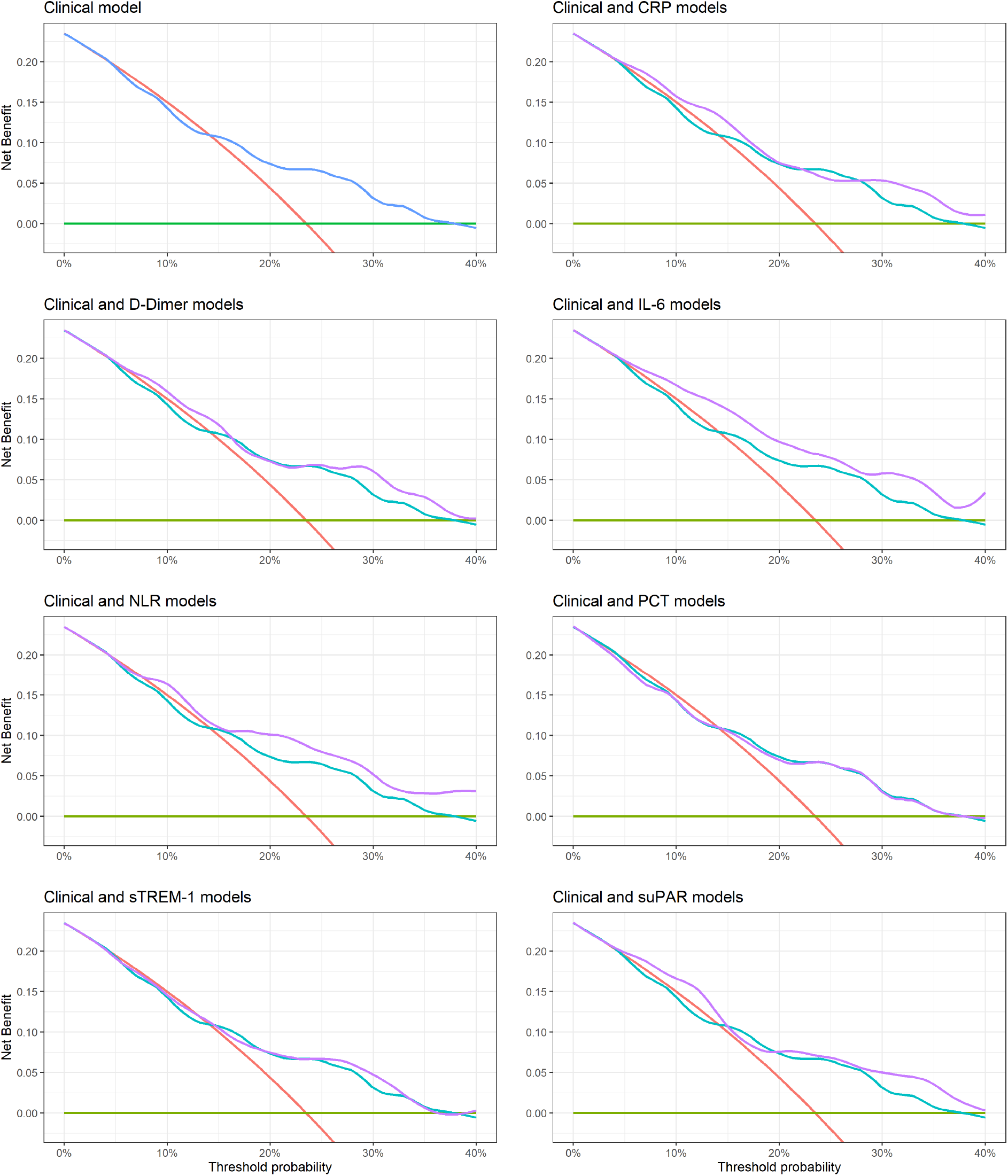
Decision curve analysis for each model in the validation cohort. The net benefit for each model is compared to an “admit-all” (red line) and “admit-none” (green line) approach, and each model containing a biochemical biomarker (purple line) is also compared to the model containing only clinical variables (blue line). A threshold probability of 5% indicates a scenario where the value of 1 TP (patient admitted who will subsequently require oxygen) is equivalent to 19 FPs (patients admitted who will not subsequently require oxygen).

## DISCUSSION

We report the development and temporal validation of three promising clinical prediction models to assist with the assessment of patients with moderate COVID-19. The models combine three simple parameters (age, sex, and SpO_2_) with measurement of a single biochemical biomarker (IL-6, NLR or suPAR), quantifiable using commercially-available near-patient tests. The models would benefit from additional validation before being recommended for use, but if these results are confirmed would be practical for use in resource-constrained settings. They could help decompress overstretched healthcare systems by supporting clinicians to identify which patients are most appropriate for community-based management.

We included patients in whom there is clinical uncertainty as to whether admission is warranted, and adopted an analytical approach which acknowledged that the trade-offs inherent in this decision will vary at different stages of the pandemic and in different healthcare settings. We used specific systemic symptoms to define moderate severity disease rather than the WHO-CPS, recognising, as did the scale’s original authors, that the lower-end of the WHO-CPS is subjective.^19^ Performance of any prediction model is sensitive to the prevalence of the outcome it aims to predict and thus we hope our more transferable study entry criteria will better standardise the outcome prevalence and facilitate model transportability; we followed the widely-used ISARIC case report form to define symptoms to permit validation by other groups.^33^

Our approach focussed on quantifying the added value of host biomarkers. We recognise that laboratory tests carry an opportunity cost, especially when resources are limited. Although a model containing clinical parameters alone would be simpler to implement, our analyses indicate that inclusion of one biomarker test would allow use of the model in a broader range of contexts, including when bed pressures are less acute early in a COVID-19 surge.

Our models have face validity. All clinical and laboratory predictors have been implicated in the pathogenesis and risk stratification of COVID-19 by previous studies.^10,17,23,25,27,29^ Similar to others, we found that age and sex were not strongly associated with risk of deterioration, in contrast to their well-recognised association with COVID-19 mortality.^23^ This underlines the importance of developing models for specific clinical use-cases. Models developed for mortality are not necessarily applicable to less severe disease, just as models developed in well-resourced healthcare systems may not generalise to LMIC settings.^34^

Reassuringly, the three biochemical biomarkers that demonstrate most promise in our study have biological plausibility. In addition to being a therapeutic target,^35^ raised IL-6 levels predict development of an oxygen requirement,^27,28^ and along with an elevated NLR, form part of the COVID-19-associated hyperinflammatory syndrome (cHIS) diagnostic criteria.^36^ Elevated suPAR levels are associated with disease severity and progression in both moderate and severe COVID-19,^29,37^ and have been used for stratification into trials of immunomodulatory agents.^38^ Just as no silver bullet exists for any problem in healthcare,^39^ no single biomarker will provide optimal risk stratification for patients with COVID-19. However, our results suggest that when used in conjunction with simple clinical parameters, these biomarkers, which are measurable with commercially-available near-patient tests, could help clinicians risk stratify patients with moderate COVID-19.

We addressed the limitations identified in other COVID-19 prognostic models by following the TRIPOD guidelines,^18^ and using a prospectively collected dataset with minimal loss-to-follow-up and missing data.^8^ Nevertheless, the small validation cohort (determined by the natural history of the pandemic in India) limits our ability to draw strong conclusions. Although the same models appeared superior in the different analyses we performed, further external validation is required before they can be recommended for use; the PRIORITISE study continues to recruit in Brazil and we have published our full model to encourage independent validation.

No vaccinated individuals were included in the study. The models may require recalibration for use in vaccinated populations with lower baseline risk of progression to severe COVID-19. However, it is important to note that only 15/54 African countries met the WHO target of vaccinating 10% of their population by the end of September 2021.^40^ An estimated 55-70% vaccination coverage is required to achieve herd immunity for a vaccine with 90% efficacy.^41^ Unfortunately, the timelines for adequate vaccination coverage in many LMICs are likely to be long.

The models were developed and validated within the Indian healthcare context. Whilst the two sites are over 2,000 kilometres apart, data from another continent would be important to assess generalisability. Although there are no licensed treatments for moderate COVID-19 available in LMIC contexts, it is possible that immunomodulatory therapies that have shown benefit in later-stage disease may provide benefit in a subset of moderate patients with particular immunological signatures.^35,38,42,43^ In our context, corticosteroids were readily available and often self-prescribed or used off-license. Although steroid use was associated with some candidate predictors, it was not associated with the primary outcome and is therefore unlikely to have confounded the observed association (appendix p19-20).

We selected oxygen requirement as our primary outcome as this reflects a clinically meaningful endpoint and the practical ceiling of care in many LMIC settings. We opted to use an SpO_2_/FiO_2_ < 400 for participants without documented hypoxia or tachypnoea prior to initiation of supplemental oxygen, as the threshold for oxygen therapy can be subjective and vary depending on available resources.^19,22^ It is unlikely that our outcome lacked sensitivity; only one participant who received supplemental oxygen did not meet the primary outcome. It may have lacked specificity (12 participants who met the primary outcome did not receive supplemental oxygen and calculation of FiO_2_ in non-ventilated patients can overestimate pulmonary dysfunction),^44^ but sensitivity would always be prioritised in a tool to inform community-based management. Furthermore, any outcome misclassification is likely to have reduced, rather than exaggerated, the prognostic performance of the candidate predictors and models.^45^

We did not explore the roles of the virus and host in influencing the risk of deterioration, as respiratory specimens were collected using different techniques and assayed on different PCR platforms at each site. However, when stratified by collection technique and platform, Ct value was not associated with the risk of deterioration (appendix p21). In keeping with others, we found that seronegativity at enrolment was associated with an increased risk of deterioration (49/190 [25·8%] vs. 37/222 [16·7%]; X^2^ = 5.16; p = 0·023).^43,46^ As near-patient antibody tests are available this warrants further exploration, acknowledging that this is likely most relevant in patients without a history of previous COVID-19 illness or vaccination.

In conclusion, we present three clinical prediction models that could help clinicians to identify patients with moderate COVID-19 who are suitable for community-based management. The models address an unmet need in the COVID-19 care continuum. They are of particular relevance where resources are scarce and, if validated, would be practical for implementation in many LMIC contexts. Routinely collected data from MSF medical facilities across 26 LMICs indicate that 54·4% (18,400 / 33,780) of patients presenting with clinically-suspected COVID-19 between March 2020 and November 2021 whom might be considered for admission (moderate, severe or critical disease), or 16·2% of all patients (18,400 / 113,455), would have been eligible for assessment using our models, illustrating the potential for widespread impact.

## Data Availability

De-identified, individual participant data from this study will be available to researchers whose proposed purpose of use is approved by the data access committees at M&eacutedecins Sans Fronti&egraveres and the Mahidol-Oxford Tropical Medicine Research Unit. Inquiries or requests for the data may be sent to. Researchers interested in accessing biobanked samples should contact the corresponding authors who will coordinate with the respective institutions.

https://osf.io/dxq43/

## CONTRIBUTORS

AC, ND, AD, CJ, PT, GMV, YL, and SB conceptualised the study. ATLC conducted the biomarker scoping review. AC, SD, JO, YL, and SB shortlisted the candidate biomarkers. PG, KG, DB, RK, SK, AL, AM, VeKS, JS, and PK collected the clinical data. TG, ShK, VK, DK, MM, MRG, SaS, ViKS, and SB were responsible for specimen processing and the laboratory assays. AC, RM, PG, LM, SRG, SH, CM, and NW curated the data. AC, LM, CK, and SB wrote and approved the statistical analysis plan. AC, RM, LM, and CK did the formal analysis. CM, JT, and NW were responsible for study monitoring. AC wrote the original draft of the manuscript. AC, RM, PG, KG, DB, LM, ATLC, ND, SD, AD, TG, SRG, SH, CJ, RK, SaK, ShK, VK, DK, AL, AM, CM, MM, JO, MRG, SaS, VeKS, ViKS, JS, SuS, JT, PT, AMGV, NW, PK, GMV, CK, YL, and SB reviewed, edited, and approved the manuscript. AC, LM, CK, and SB verified the underlying data.

## DATA SHARING

De-identified, individual participant data from this study will be available to researchers whose proposed purpose of use is approved by the data access committees at Médecins Sans Frontières and the Mahidol-Oxford Tropical Medicine Research Unit. Inquiries or requests for the data may be sent to. Researchers interested in accessing biobanked samples should contact the corresponding authors who will coordinate with the respective institutions.

## ACKNOWLEDGEMENTS

We are grateful to the members of the External Advisory Panel, led by Prof. Nicholas J White (MORU), for their advice and guidance throughout the study. We also acknowledge the support of Kundavaram PP Abhilash (CMC), OC Abraham (CMC), T Balamugesh (CMC), Thambu David (CMC), Divya Dayanand (CMC), Biju George (CMC), Richa Gupta (CMC), Samuel G Hansdak (CMC), Ramya Iyadurai (CMC), Rajiv Karthik (CMC), Sharwar Kazmi (MSF), Mavuto Mukaka (MORU), Sowmya Sathyendra (CMC), Merylin Sebastian (CMC), Christopher Smith (LSHTM), Ramesh Vishwakarma (MORU), Sophie Yacoub (OUCRU), and Anand Zachariah (CMC). Funding for this work was provided by MSF, India. Wellcome Trust provides core funding to the Mahidol-Oxford Tropical Medicine Research Unit in Bangkok [220211; 215604/Z/19/Z], which supported the design, monitoring and analysis of the study. CK is supported by a Wellcome Trust/Royal Society Sir Henry Dale Fellowship [211182/Z/18/Z]. For the purpose of open access, the author has applied a CC BY public copyright license to any Author Accepted Manuscript version arising from this submission.

