## Supplementary appendix for "Facilitating safe discharge through predicting disease progression in moderate COVID-19: a prospective cohort study to develop and validate a clinical prediction model in resource-limited settings"

**CONTENTS** **PAGE**


| | |
|---|---|
| **1. Results of scoping review** | 2 |
| **2. Examples of near-patient tests for candidate biomarkers** | 4 |
| **3. Proportion of missing data for candidate predictors** | 5 |
| **4. Baseline characteristics of development and validation cohorts** | 6 |
| **5. Levels of supplemental oxygen support for participants who met the primary outcome** | 10 |
| **6. Kaplan-Meier curves to indicate time to meeting the primary outcome** | 11 |
| **7. Unadjusted and adjusted associations between candidate predictors and primary outcome** | 12 |
| **8. Classification measures for the different models in the validation cohort** | 14 |
| **9. Sensitivity and specificity for the different models in the validation cohort** | 17 |
| **10. Association of corticosteroid use with primary outcome** | 18 |
| **11. Association of corticosteroid use with candidate predictors** | 19 |
| **12. Association of RT-PCR Ct values with primary outcome** | 20 |

**Results of scoping review conducted to inform biomarker longlisting for PRIORITISE study.**

*Search strategy*

Date performed: 1 June 2020

("severe acute respiratory syndrome coronavirus 2"[Supplementary Concept] OR coronavirus*[tiab] OR SARS-CoV-2[tiab] OR 2019-nCoV[tiab] OR COVID-19[Supplementary Concept] OR COVID*[tiab] OR "Wuhan seafood market pneumonia"[tiab])

**AND**

(Biomarkers[MeSH] OR Biomarker*[tiab] OR "Biologic marker*"[tiab] OR "Biological marker*"[tiab] OR "Laboratory marker*"[tiab] OR "Lab marker*"[tiab]

**OR**

Prognosis[MeSH] OR Prognos*[tiab] OR ((Predict*[tiab] OR Risk*[tiab]) AND "Severity"[tiab] OR Outcome*[tiab] OR Progression[tiab] OR Deteriorat*[tiab])))

**AND**

2019/11[PDAT] : 9999[PDAT]

*Screening of results*

**Supplementary Figure 1. Screening of articles for inclusion.**

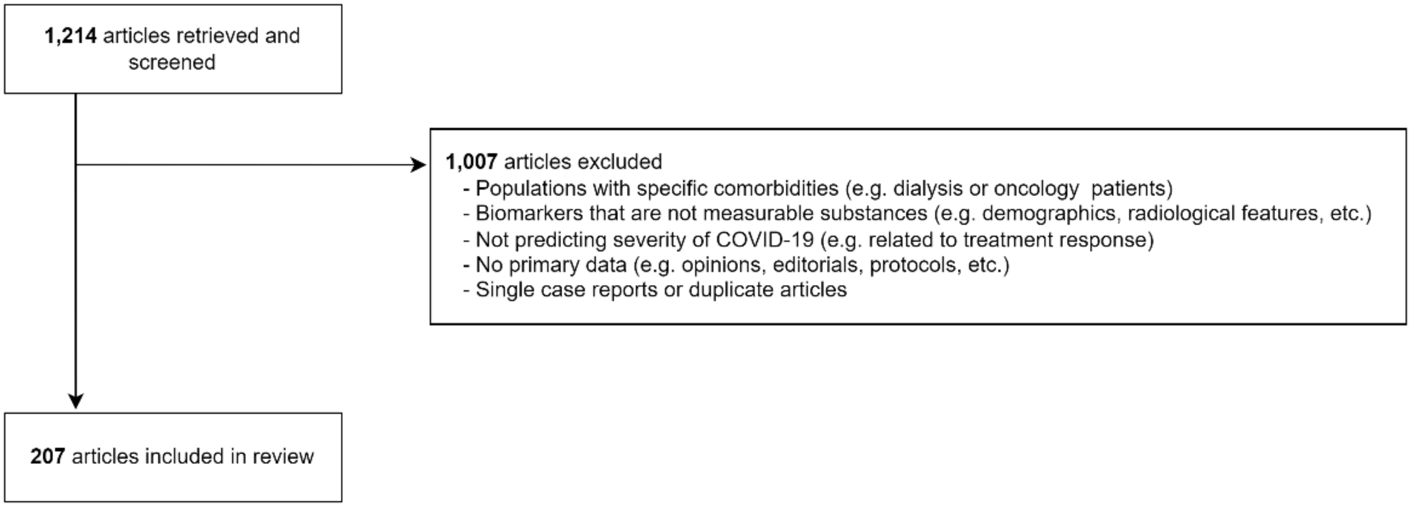

*Summary of main results*

**Supplementary Figure 2. Proportion of articles reporting statistically significant findings for different biochemical biomarkers in the prognostication of COVID-19.** Overall results (dark blue; n = 207) were broadly similar across different subgroups, including prospective studies (green; n = 16), studies outside China (red; n = 55); PCR/NGS-confirmed SARS-CoV-2 infection (yellow; n = 121); outcome determined after biomarker measurement i.e. prognostic vs. diagnostic (grey; n = 100), and studies with adjusted estimates (light blue; n = 81).

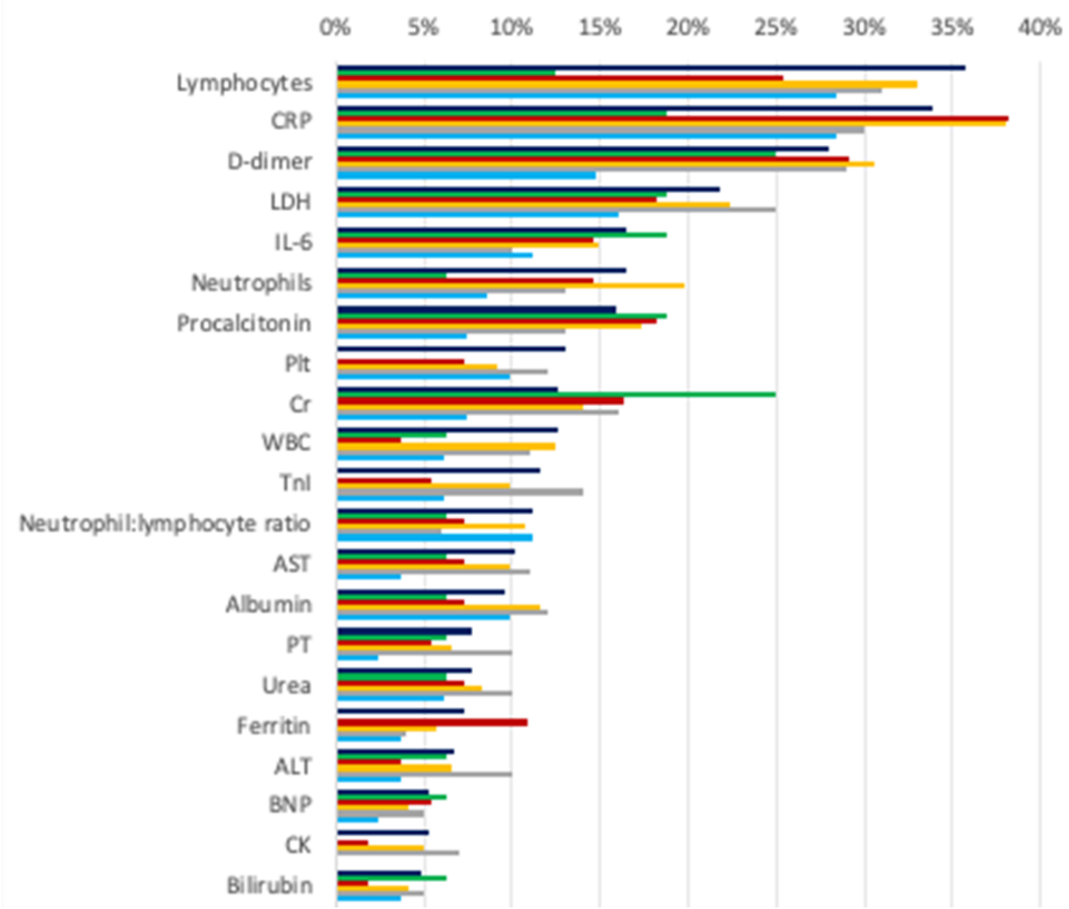

**Supplementary Table 1. Examples of commercially-available or late-stage development near-patient quantitative tests for candidate biochemical biomarkers shortlisted for inclusion in the clinical prediction models.**

| Biomarker | Manufacturer | Test characteristics | | |
|---|---|---|---|---|
| | | Sample type | Sample volume | Turn-around-time |
| CRP | NycoCard™, Abbott | Whole blood, serum or plasma | 5uL | 3 minutes |
| D-dimer | RAMP, Response Biomedical | Whole blood | 75uL | 15 minutes |
| IL-6 | IL-6, Hotgen | Serum | 100uL | 15 minutes |
| NLR | WBC DIFF, HemoCue® | Whole blood | 10uL | 5 minutes |
| PCT | BRAHMS PCT direct, Roche | Whole blood | 20uL | 20 minutes |
| sTREM-1 | FIND, personal communication | Whole blood | TBC | TBC |
| suPAR | suPARnostic®, Virogates | Plasma | 10uL | 20 minutes |

**Supplementary Table 2. Number of participants with missing data for candidate predictor variables, stratified by cohort.**

| Variable | Number missing | | |
|---|---|---|---|
| | **Development cohort** (n = 257) | **Validation cohort** (n = 166) | **Overall** (n = 423) |
| Age (years) | 0/257 | 0/166 | **0/423** |
| Sex | 0/257 | 0/166 | **0/423** |
| $SpO_2$ | 0/257 | 0/166 | **0/423** |
| CRP (mg/l) | 0/257 | 8/166 (4·8%) | **8/423 (1·9%)** |
| D-dimer (ng/ml) | 1/257 (0·4%) | 2/166 (1·2%) | **3/423 (0·7%)** |
| IL-6 (pg/ml) | 0/257 | 2/166 (1·2%) | **2/423 (0·5%)** |
| NLR | 10/257 (3·9%) | 1/166 (0·6%) | **11/423 (2·6%)** |
| PCT (ng/ml) | 0/257 | 2/166 (1·2%) | **2/423 (0·5%)** |
| sTREM-1 (pg/ml) | 0/257 | 2/166 (1·2%) | **2/423 (0·5%)** |
| suPAR (ng/ml) | 0/257 | 0/166 | **0/423** |

**Supplementary Table 3. Baseline characteristics of development and validation cohorts, stratified by primary outcome status, including additional details on presenting symptoms and reported comorbidities.** Details for presenting symptoms and reported comorbidities with prevalence ≥ 5% in any of the outcome groups for either the development or validation cohorts are reported. Data on other reported comorbidities (chronic neurological disorder, liver disease, active tuberculosis, malignant neoplasm, HIV, and other immunosuppression) and presenting symptoms (otalgia, wheeze, lower chest indrawing, altered consciousness, seizures, conjunctivitis, skin rash, skin ulcers, and lymphadenopathy) not reported as prevalence < 5% in all outcome groups. *Missing data for continuous variables: BMI = 1; CRP = 8, D-dimer = 3, IL-6 = 2, NLR = 12; PCT = 2; sTREM-1 = 2; Ct value = 181. §Different specimen collection procedures and PCR assays were used at each site (appendix p21). †Seronegative defined as negative for both SARS-CoV-2 IgG and IgM antibodies. Median values (IQR) and p-values for Wilcoxon rank sum tests are reported for continuous variables. Fisher's exact or Pearson's Chi-squared test p-values are reported for categorical variables.

| Baseline characteristic | DEVELOPMENT COHORT | | | | VALIDATION COHORT | | | | p-value |
|---|---|---|---|---|---|---|---|---|---|
| | Overall (n = 257) | Developed oxygen requirement | | p-value | Overall (n = 166) | Developed oxygen requirement | | p-value | |
| | | No (n = 207) | Yes (n = 50) | | | No (n = 127) | Yes (n = 39) | | |
| **Demographics** | | | | | | | | | |
| Age (years) | 52·0 (40·0 to 61·0) | 52·0 (40·0 to 60·0) | 54·0 (42·2 to 62·0) | 0·3 | 54·0 (41·2 to 63·0) | 55·0 (41·5 to 63·0) | 54·0 (41·0 to 66·0) | 0·8 | 0·2 |
| Male sex | 185 / 257 (72%) | 144 / 207 (70%) | 41 / 50 (82%) | 0·079 | 101 / 166 (61%) | 76 / 127 (60%) | 25 / 39 (64%) | 0·6 | 0·017 |
| BMI (kg/m²) * | 26·0 (23·6 to 28·7) | 26·2 (23·8 to 28·8) | 25·8 (22·4 to 28·3) | 0·4 | 24·9 (23·4 to 27·6) | 24·8 (23·4 to 27·6) | 26·1 (23·7 to 27·6) | 0·7 | 0·071 |
| **Vital signs** | | | | | | | | | |
| Heart rate (bpm) | 88·0 (80·0 to 97·0) | 86·0 (79·0 to 96·0) | 90·0 (86·0 to 99·5) | 0·010 | 84·0 (74·0 to 92·0) | 84·0 (74·0 to 90·0) | 84·0 (77·0 to 94·0) | 0·4 | < 0·001 |
| Respiratory rate (bpm) | 22·0 (22·0 to 24·0) | 22·0 (22·0 to 24·0) | 22·0 (22·0 to 24·0) | 0·3 | 24·0 (22·0 to 24·0) | 22·0 (22·0 to 24·0) | 24·0 (22·0 to 24·0) | 0·12 | 0·006 |
| Oxygen saturation (%) | 98·0 (96·0 to 99·0) | 98·0 (97·0 to 99·0) | 96·0 (95·2 to 98·0) | < 0·001 | 98·0 (96·0 to 99·0) | 98·0 (96·0 to 99·0) | 96·0 (95·5 to 98·0) | 0·002 | 0·5 |
| Axillary temperature (°C) | 36·8 (36·4 to 37·1) | 36·7 (36·4 to 37·0) | 36·9 (36·5 to 37·2) | 0·045 | 36·9 (36·7 to 37·2) | 36·9 (36·7 to 37·2) | 37·0 (36·9 to 37·2) | 0·053 | < 0·001 |

| | | | | | | | | | |
|---|---|---|---|---|---|---|---|---|---|
| Systolic BP (mmHg) | 128·0 (116·0 to 138·0) | 128·0 (116·0 to 140·0) | 126·0 (118·0 to 134·8) | 0·7 | 121·0 (110·0 to 130·0) | 120·0 (110·0 to 130·0) | 122·0 (110·0 to 131·0) | > 0·9 | < 0·001 |
| Diastolic BP (mmHg) | 80·0 (72·0 to 88·0) | 80·0 (72·0 to 88·0) | 79·0 (70·0 to 88·0) | 0·3 | 76·0 (70·0 to 82·0) | 76·0 (70·0 to 82·0) | 74·0 (67·0 to 80·0) | 0·3 | < 0·001 |
| qSOFA score ≥ 2 | 13 / 257 (5·1%) | 9 / 207 (4·3%) | 4 / 50 (8·0%) | 0·3 | 16 / 166 (9·6%) | 10 / 127 (7·9%) | 6 / 39 (15%) | 0·2 | 0·069 |
| **Comorbidities** | | | | | | | | | |
| Current smokers | 10 / 257 (3·9%) | 8 / 207 (3·9%) | 2 / 50 (4·0%) | > 0·9 | 4 / 166 (2·4%) | 3 / 127 (2·4%) | 1 / 39 (2·6%) | > 0·9 | 0·4 |
| Reported comorbidity [#] | 165 / 257 (64%) | 128 / 207 (62%) | 37 / 50 (74%) | 0·11 | 117 / 166 (70%) | 91 / 127 (72%) | 26 / 39 (67%) | 0·6 | 0·2 |
| Cardiovascular disease | 21 / 252 (8·3%) | 16 / 203 (7·9%) | 5 / 49 (10%) | 0·6 | 19 / 166 (11%) | 15 / 127 (12%) | 4 / 39 (10%) | > 0·9 | 0·3 |
| Diabetes | 92 / 255 (36%) | 73 / 205 (36%) | 19 / 50 (38%) | 0·8 | 64 / 166 (39%) | 55 / 127 (43%) | 9 / 39 (23%) | 0·023 | 0·6 |
| Hypertension | 89 / 256 (35%) | 73 / 206 (35%) | 16 / 50 (32%) | 0·6 | 64 / 166 (39%) | 47 / 127 (37%) | 17 / 39 (44%) | 0·5 | 0·4 |
| Chronic lung disease | 3 / 252 (1·2%) | 3 / 203 (1·5%) | 0 / 49 (0%) | > 0·9 | 3 / 166 (1·8%) | 1 / 127 (0·8%) | 2 / 39 (5·1%) | 0·14 | 0·7 |
| Asthma | 16 / 252 (6·3%) | 13 / 203 (6·4%) | 3 / 49 (6·1%) | > 0·9 | 11 / 166 (6·6%) | 9 / 127 (7·1%) | 2 / 39 (5·1%) | > 0·9 | > 0·9 |
| Chronic kidney disease | 10 / 252 (4·0%) | 8 / 203 (3·9%) | 2 / 49 (4·1%) | > 0·9 | 9 / 166 (5·4%) | 5 / 127 (3·9%) | 4 / 39 (10%) | 0·2 | 0·5 |
| **Presenting illness** | | | | | | | | | |
| Symptom duration (days) | 6·0 (4·0 to 8·0) | 6·0 (4·0 to 8·0) | 5·5 (5·0 to 7·0) | 0·8 | 6·0 (4·0 to 8·0) | 6·0 (3·5 to 8·0) | 5·0 (4·0 to 7·0) | 0·5 | 0·10 |
| History of fever | 243 / 257 (95%) | 196 / 207 (95%) | 47 / 50 (94%) | 0·7 | 155 / 166 (93%) | 118 / 127 (93%) | 37 / 39 (95%) | > 0·9 | 0·6 |
| Breathlessness | 154 / 257 (60%) | 119 / 207 (57%) | 35 / 50 (70%) | 0·11 | 90 / 166 (54%) | 65 / 127 (51%) | 25 / 39 (64%) | 0·2 | 0·2 |

| | | | | | | | | | |
|---|---|---|---|---|---|---|---|---|---|
| Chest pain | 59 / 257 (23%) | 48 / 207 (23%) | 11 / 50 (22%) | 0·9 | 15 / 166 (9·0%) | 9 / 127 (7·1%) | 6 / 39 (15%) | 0·12 | < 0·001 |
| Abdominal pain | 35 / 257 (14%) | 32 / 207 (15%) | 3 / 50 (6·0%) | 0·080 | 15 / 166 (9·0%) | 12 / 127 (9·4%) | 3 / 39 (7·7%) | > 0·9 | 0·2 |
| Diarrhoea | 80 / 257 (31%) | 65 / 207 (31%) | 15 / 50 (30%) | 0·8 | 47 / 166 (28%) | 33 / 127 (26%) | 14 / 39 (36%) | 0·2 | 0·5 |
| Severe myalgia | 140 / 257 (54%) | 110 / 207 (53%) | 30 / 50 (60%) | 0·4 | 75 / 166 (45%) | 65 / 127 (51%) | 10 / 39 (26%) | 0·005 | 0·062 |
| Sore throat | 87 / 257 (34%) | 69 / 207 (33%) | 18 / 50 (36%) | 0·7 | 31 / 166 (19%) | 23 / 127 (18%) | 8 / 39 (21%) | 0·7 | < 0·001 |
| Rhinorrhoea | 49 / 257 (19%) | 42 / 207 (20%) | 7 / 50 (14%) | 0·3 | 19 / 166 (11%) | 14 / 127 (11%) | 5 / 39 (13%) | 0·8 | 0·037 |
| Cough | 205 / 257 (80%) | 160 / 207 (77%) | 45 / 50 (90%) | 0·045 | 144 / 166 (87%) | 109 / 127 (86%) | 35 / 39 (90%) | 0·5 | 0·065 |
| Arthralgia | 49 / 257 (19%) | 37 / 207 (18%) | 12 / 50 (24%) | > 0·9 | 2 / 166 (1·2%) | 2 / 127 (1·6%) | 0 / 39 (0%) | > 0·9 | < 0·001 |
| Fatigue | 190 / 257 (74%) | 153 / 207 (74%) | 37 / 50 (74%) | > 0·9 | 97 / 166 (58%) | 76 / 127 (60%) | 21 / 39 (54%) | 0·5 | < 0·001 |
| Anorexia | 69 / 257 (27%) | 52 / 207 (25%) | 17 / 50 (34%) | 0·2 | 14 / 166 (8·4%) | 12 / 127 (9·4%) | 2 / 39 (5·1%) | 0·5 | < 0·001 |
| Nausea or vomiting | 55 / 257 (21%) | 42 / 207 (20%) | 13 / 50 (26%) | 0·4 | 23 / 166 (14%) | 17 / 127 (13%) | 6 / 39 (15%) | 0·8 | 0·051 |
| Headache | 75 / 257 (29%) | 65 / 207 (31%) | 10 / 50 (20%) | 0·11 | 41 / 166 (25%) | 32 / 127 (25%) | 9 / 39 (23%) | 0·8 | 0·3 |
| Anosmia | 60 / 257 (23%) | 52 / 207 (25%) | 8 / 50 (16%) | 0·2 | 13 / 166 (7·8%) | 9 / 127 (7·1%) | 4 / 39 (10%) | 0·5 | < 0·001 |
| Aguesia | 60 / 257 (23%) | 51 / 207 (25%) | 9 / 50 (18%) | 0·3 | 19 / 166 (11%) | 14 / 127 (11%) | 5 / 39 (13%) | 0·8 | 0·002 |
| **Host biomarkers** | | | | | | | | | |
| CRP (mg/l) * | 24·4 (3·9 to 88·9) | 17·9 (2·8 to 85·4) | 62·5 (19·7 to 134·4) | < 0·001 | 58·1 (17·2 to 147·1) | 42·5 (12·3 to 111·9) | 95·8 (52·8 to 176·9) | < 0·001 | < 0·001 |

| | | | | | | | | | |
|---|---|---|---|---|---|---|---|---|---|
| D-dimer (ng/ml) * | 725·0 (382·4 to 1,466·4) | 640·6 (329·7 to 1,234·9) | 1,201·7 (679·9 to 2,307·0) | < 0·001 | 968·2 (620·7 to 1,599·0) | 918·8 (579·0 to 1,454·9) | 1,148·1 (829·5 to 3,200·2) | 0·009 | < 0·001 |
| IL-6 (pg/ml) * | 11·0 (4·9 to 36·2) | 8·7 (4·2 to 27·9) | 36·4 (18·4 to 70·7) | < 0·001 | 31·6 (13·9 to 63·0) | 24·4 (11·4 to 47·2) | 71·1 (39·4 to 98·9) | < 0·001 | < 0·001 |
| NLR * | 3·2 (1·9 to 4·9) | 2·9 (1·7 to 4·5) | 4·4 (3·2 to 7·2) | < 0·001 | 2·8 (1·8 to 5·4) | 2·5 (1·6 to 4·2) | 5·3 (2·7 to 7·0) | < 0·001 | 0·6 |
| PCT (ng/ml) * | 0·1 (0·1 to 0·2) | 0·1 (0·1 to 0·1) | 0·1 (0·1 to 0·2) | < 0·001 | 0·1 (0·1 to 0·2) | 0·1 (0·1 to 0·2) | 0·1 (0·1 to 0·3) | 0·3 | 0·004 |
| sTREM-1 (pg/ml) * | 378·0 (265·0 to 537·0) | 362·0 (259·0 to 522·0) | 424·5 (306·8 to 649·5) | 0·055 | 419·0 (285·0 to 596·8) | 389·0 (282·0 to 562·0) | 437·0 (349·0 to 660·8) | 0·13 | 0·2 |
| suPAR (ng/ml) | 4·2 (3·1 to 5·8) | 4·0 (2·9 to 5·5) | 5·4 (4·0 to 6·8) | < 0·001 | 4·1 (3·1 to 5·6) | 3·8 (2·9 to 5·1) | 5·5 (3·9 to 6·7) | < 0·001 | 0·9 |
| **Viral markers** | | | | | | | | | |
| Ct value *§ | 26·0 (20·7 to 30·8) | 26·0 (20·6 to 30·1) | 26·4 (22·0 to 31·4) | 0·6 | 32·1 (28·3 to 36·2) | 32·8 (28·1 to 36·2) | 31·5 (28·4 to 36·0) | > 0·9 | < 0·001 |
| Seronegative † | 117 / 252 (46%) | 90 / 203 (44%) | 27 / 49 (55%) | 0·2 | 73 / 160 (46%) | 51 / 123 (41%) | 22 / 37 (59%) | 0·054 | 0·9 |

**Supplementary Table 4. Maximum level of supplemental oxygen support received by participants who met the primary outcome, stratified by cohort.** One participant (not included in this table) received supplemental oxygen via nasal cannula but did not meet the primary outcome as their $SpO_2/FiO_2$ remained above 400. *One participant met the endpoint on the basis of being prescribed oxygen at another licensed medical facility; all other participants who met the endpoint did so on the basis of an $SpO_2 < 94\%$ and/or $SpO_2/FiO_2 < 400$ and/or death.

| | **Development** (n = 257) | **Validation** (n = 166) | **Overall** (n=423) |
|---|---|---|---|
| **Number of participants meeting primary outcome** | 50 | 39 | 89 |
| **Deaths** | 2 | 9 | 11 |
| **Mechanical ventilation** | 1 | 1 | 2 |
| **Non-invasive ventilation** | 5 | 10 | 15 |
| **Supplemental oxygen via face mask and/or nasal cannula*** | 32 | 17 | 49 |
| **No supplemental oxygen received** | 10 | 2 | 12 |

**Supplementary Figure 3. Kaplan-Meier survival curves indicating the time to meeting the primary outcome, stratified by cohort.** Coloured lines indicate survival probability for each cohort and shaded areas indicate 95% confidence intervals.

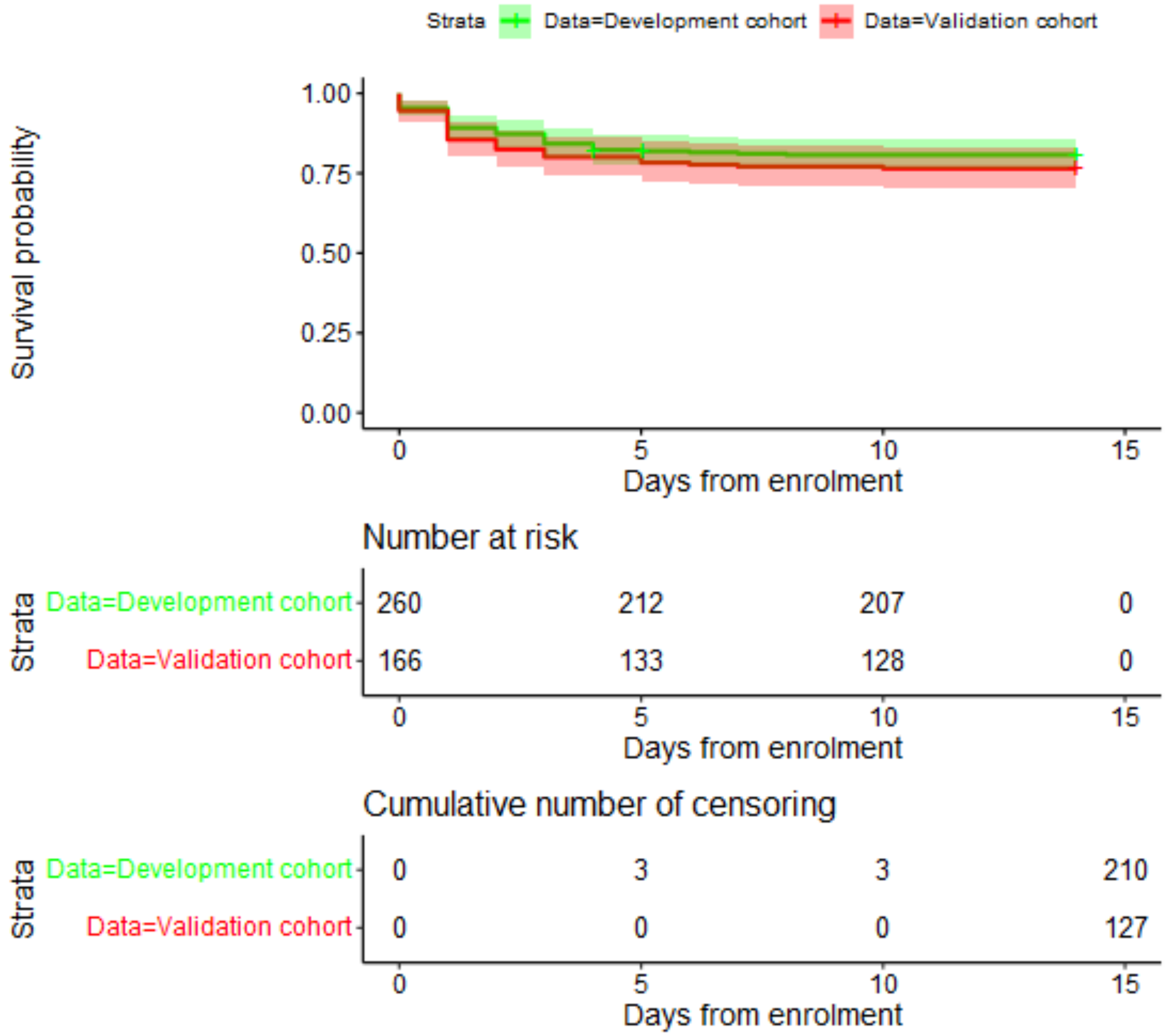

**Supplementary Table 5. Unadjusted and adjusted associations between candidate predictors and primary outcome in each model in the development cohort**. 1,000 bootstrap samples were used to estimate 95% confidence intervals for the Ridge regression coefficients. * Transformations used due to non-linear relationship between predictor and outcome: $Log_{10}$ used for CRP and D-dimer; $\frac{1}{\sqrt{IL-6/100}}$ used for IL-6. As a result, odds ratios are not comparable between continuous predictors, as scales cannot be standardised. Odds ratios are expressed for a one unit increase in non-transformed continuous predictors (age, NLR, PCT, sTREM-1, suPAR) and a ten-fold increase in CRP or D-dimer.

| Variable | Unadjusted odds ratio (95% CI) | Unadjusted c-statistic (95% CI) | Ridge regression coefficient (95% CI) | Adjusted odds ratio (95% CI) |
|---|---|---|---|---|
| **Clinical model** | | | | |
| Intercept | NA | NA | 41·89 (23·14 to 67·65) | NA |
| Male sex | 1·99 (0·98 to 5·24) | NA | 0·31 (-0·30 to 1·22) | 1·37 (0·74 to 3·37) |
| Age (years) | 1·01 (0·99 to 1·03) | 0·55 (0·48 to 0·64) | 0·00 (-0·02 to 0·02) | 1·00 (0·98 to 1·02) |
| $SpO_2$ | **0·56 (0·43 to 0·71)** | 0·72 (0·64 to 0·80) | -0·45 (-0·71 to -0·26) | **0·64 (0·49 to 0·77)** |
| **CRP model** | | | | |
| Intercept | NA | NA | 40·47 (21·51 to 66·25) | NA |
| Male sex | NA | NA | 0·16 (-0·56 to 1·03) | 1·18 (0·57 to 2·79) |
| Age (years) | NA | NA | -0·00 (-0·03 to 0·02) | 1·00 (0·97 to 1·02) |
| $SpO_2$ | NA | NA | -0·44 (-0·71 to -0·24) | **0·64 (0·49 to 0·78)** |
| CRP (mg/l) * | **2·24 (1·59 to 3·52)** | 0·66 (0·58 to 0·73) | 0·54 (0·23 to 0·98) | **1·71 (1·26 to 2·65)** |
| **D-dimer model** | | | | |
| Intercept | NA | NA | 38·15 (22·19 to 67·18) | NA |
| Male sex | NA | NA | 0·34 (-0·29 to 1·26) | 1·40 (0·75 to 3·54) |
| Age (years) | NA | NA | -0·00 (-0·03 to 0·02) | 1·00 (0·97 to 1·02) |
| $SpO_2$ | NA | NA | -0·44 (-0·75 to -0·28) | **0·65 (0·47 to 0·76)** |
| D-dimer (ng/ml) * | **3·32 (1·85 to 6·89)** | 0·67 (0·60 to 0·75) | 0·98 (0·47 to 1·88) | **2·67 (1·60 to 6·54)** |
| **IL-6 model** | | | | |
| Intercept | NA | NA | 39·92 (24·84 to 64·91) | NA |
| Male sex | NA | NA | 0·11 (-0·64 to 1·02) | 1·11 (0·53 to 2·76) |
| Age (years) | NA | NA | -0·01 (-0·03 to 0·02) | 0·99 (0·97 to 1·02) |

| | | | | |
|---|---|---|---|---|
| $SpO_2$ | NA | NA | -0·41 (-0·68 to -0·25) | **0·66 (0·51 to 0·78)** |
| IL-6 (pg/ml) * | **0·53 (0·39 to 0·65)** | 0·75 (0·68 to 0·82) | -0·44 (-0·66 to -0·25) | **0·65 (0·52 to 0·78)** |
| **NLR model** | | | | |
| Intercept | NA | NA | 38·27 (19·93 to 66·20) | NA |
| Male sex | NA | NA | 0·21 (-0·45 to 0·98) | 1·24 (0·63 to 2·65) |
| Age (years) | NA | NA | -0·00 (-0·03 to 0·02) | 1·00 (0·97 to 1·02) |
| $SpO_2$ | NA | NA | -0·41 (-0·71 to -0·22) | **0·66 (0·49 to 0·80)** |
| NLR | **1·15 (1·07 to 1·27)** | 0·69 (0·61 to 0·77) | 0·10 (0·04 to 0·20) | **1·10 (1·04 to 1·22)** |
| **PCT model** | | | | |
| Intercept | NA | NA | 41·03 (1·03 to 66·59) | NA |
| Male sex | NA | NA | 0·32 (-0·29 to 1·19) | 1·37 (0·75 to 3·28) |
| Age (years) | NA | NA | 0·00 (-0·02 to 0·02) | 1·00 (0·98 to 1·02) |
| $SpO_2$ | NA | NA | -0·44 (-0·71 to -0·03) | **0·64 (0·49 to 0·97)** |
| PCT (ng/ml) | 0·96 (0·80 to 1·74) | 0·66 (0·58 to 0·74) | -0·03 (-0·03 to 1·28) | 0·97 (0·97 to 3·60) |
| **sTREM-1 model** | | | | |
| Intercept | NA | NA | 42·55 (22·26 to 70·09) | NA |
| Male sex | NA | NA | 0·28 (-0·35 to 1·11) | 1·33 (0·71 to 3·02) |
| Age (years) | NA | NA | 0·00 (-0·02 to 0·02) | 1·00 (0·98 to 1·02) |
| $SpO_2$ | NA | NA | -0·46 (-0·746 to -0·25) | **0·63 (0·48 to 0·78)** |
| sTREM-1 (pg/ml) | 1·00 (1·00 to 1·00) | 0·58 (0·48 to 0·68) | 0·00 (-0·00 to 0·00) | 1·00 (1·00 to 1·00) |
| **suPAR model** | | | | |
| Intercept | NA | NA | 38·82 (21·24 to 66·61) | NA |
| Male sex | NA | NA | 0·38 (-0·24 to 1·32) | 1·46 (0·79 to 3·74) |
| Age (years) | NA | NA | -0·00 (-0·03 to 0·02) | 1·00 (0·98 to 1·02) |
| $SpO_2$ | NA | NA | -0·42 (-0·71 to -0·24) | **0·65 (0·49 to 0·79)** |
| suPAR (ng/ml) | **1·20 (1·11 to 1·36)** | 0·68 (0·61 to 0·76) | 0·14 (0·06 to 0·25) | **1·15 (1·06 to 1·29)** |

**Supplementary Table 6. Sensitivity, specificity, negative predictive value and positive predictive value for each model at different cut-offs in the validation cohort**. A cut-off of 0·1 reflects a management strategy in which any patient with a predicted risk of requiring oxygen ≥ 10% is admitted.

| | Sensitivity (95% CI) | Specificity (95% CI) | Negative likelihood ratio (95% CI) | Positive likelihood ratio (95% CI) | Negative predictive value (95% CI) | Positive predictive value (95% CI) |
|---|---|---|---|---|---|---|
| **Clinical model** | | | | | | |
| 0·1 | 89·7 (75·8 to 97·1) | 25·2 (17·9 to 33·7) | 0·41 (0·15 to 1·08) | 1·20 (1·04 to 1·39) | 88·9 (73·9 to 96·9) | 26·9 (19·5 to 35·4) |
| 0·15 | 76·9 (60·7 to 88·9) | 40·9 (32·3 to 50·2) | 0·56 (0·31 to 1·04) | 1·30 (1·04 to 1·63) | 85·3 (73·8 to 93·0) | 28·6 (20·2 to 38·2) |
| 0·20 | 61·5 (44·6 to 76·6) | 62·9 (53·9 to 71·4) | 0·61 (0·40 to 0·93) | 1·66 (1·19 to 2·33) | 84·2 (75·3 to 90·9) | 33·8 (23·0 to 46·0) |
| **IL-6 model** | | | | | | |
| 0·1 | 100 (90·9 to 100) | 21·3 (14·5 to 29·4) | 0 (NA) | 1·27 (1·16 to 1·39) | 100 (87·2 to 100) | 28·1 (20·8 to 36·3) |
| 0·15 | 92·3 (79·1 to 98·4) | 36·2 (27·9 to 45·2) | 0·21 (0·07 to 0·65) | 1·45 (1·23 to 1·70) | 93·9 (83·1 to 98·7) | 30·8 (22·6 to 39·9) |
| 0·20 | 82·1 (66·5 to 92·5) | 51·2 (42·2 to 60·2) | 0·35 (0·18 to 0·70) | 1·68 (1·33 to 2·12) | 90·3 (80·9 to 96·0) | 34·0 (24·6 to 44·5) |
| **NLR model** | | | | | | |
| 0·1 | 95·0 (82·7 to 99·3) | 29·9 (22·1 to 38·7) | 0·17 (0·04 to 0·68) | 1·35 (1·18 to 1·55) | 95·0 (83·1 to 99·4) | 29·4 (21·6 to 38·1) |
| 0·15 | 74·4 (57·9 to 86·9) | 49·6 (40·6 to 58·6) | 0·52 (0·29 to 0·91) | 1·48 (1·15 to 1·90) | 86·3 (76·3 to 93·2) | 31·2 (21·9 to 41·6) |
| 0·20 | 66·7 (49·8 to 80·9) | 67·7 (58·9 to 75·7) | 0·49 (0·31 to 0·78) | 2·10 (1·48 to 2·89) | 86·9 (78·6 to 92·8) | 38·8 (27·1 to 51·5) |
| **suPAR model** | | | | | | |
| 0·1 | 95·0 (82·7 to 99·4) | 33·1 (24·9 to 41·9) | 0·16 (0·04 to 0·61) | 1·42 (1·23 to 1·63) | 95·4 (84·5 to 99·4) | 30·3 (22·3 to 39·3) |
| 0·15 | 69·2 (52·4 to 82·9) | 55·9 (46·8 to 64·7) | 0·55 (0·34 to 0·90) | 1·57 (1·18 to 2·10) | 85·5 (76·1 to 92·3) | 32·5 (22·7 to 43·7) |
| 0·20 | 56·4 (39·6 to 72·2) | 70·9 (62·2 to 78·6) | 0·62 (0·42 to 0·89) | 1·94 (1·32 to 2·85) | 84·1 (75·8 to 90·4) | 37·3 (25·0 to 50·9) |
| **CRP model** | | | | | | |
| 0·1 | 92·3 (79·1 to 98·4) | 29·9 (22·1 to 38·7) | 0·26 (0·08 to 0·79) | 1·32 (1·14 to 1·52) | 92·7 (80·1 to 98·4) | 28·8 (21·1 to 37·6) |
| 0·15 | 87·2 (72·6 to 95·7) | 44·1 (35·3 to 53·2) | 0·29 (0·13 to 0·67) | 1·56 (1·28 to 1·90) | 91·8 (81·9 to 97·3) | 32·4 (23·6 to 42·2) |
| 0·20 | 69·2 (52·4 to 82·9) | 53·5 (44·5 to 62·4) | 0·57 (0·35 to 0·95) | 1·49 (1·13 to 1·97) | 85·0 (75·3 to 92·0) | 31·4 (21·8 to 42·3) |
| **D-dimer model** | | | | | | |
| 0·1 | 92·3 (79·1 to 98·4) | 29·9 (22·1 to 38·7) | 0·26 (0·08 to 0·79) | 1·32 (1·14 to 1·52) | 92·7 (80·1 to 98·5) | 28·8 (21·1 to 37·6) |
| 0·15 | 82·1 (66·5 to 92·5) | 48·8 (39·8 to 57·8) | 0·37 (0·18 to 0·74) | 1·60 (1·28 to 2·01) | 89·9 (80·2 to 95·8) | 32·9 (23·8 to 43·3) |
| 0·20 | 64·1 (47·2 to 78·8) | 59·8 (50·8 to 68·4) | 0·6 (0·39 to 0·93) | 1·60 (1·16 to 2·19) | 84·4 (75·3 to 91·2) | 32·8 (22·5 to 44·6) |
| **PCT model** | | | | | | |

| | | | | | | |
|---|---|---|---|---|---|---|
| 0·1 | 89·7<br>(75·8 to 97·1) | 25·9<br>(18·6 to 34·5) | 0·39<br>(0·15 to 1·05) | 1·21<br>(1·05 to 1·41) | 89·2<br>(74·6 to 96·8) | 27·1<br>(19·7 to 35·7) |
| 0·15 | 76·9<br>(60·7 to 88·9) | 41·7<br>(33·1 to 50·8) | 0·55<br>(0·3 to 1·02) | 1·32<br>(1·05 to 1·66) | 85·5<br>(74·2 to 93·1) | 28·9<br>(20·4 to 38·6) |
| 0·20 | 58·9<br>(42·1 to 74·4) | 64·6<br>(55·6 to 72·6) | 0·64<br>(0·43 to 0·95) | 1·66<br>(1·17 to 2·37) | 83·7<br>(78·8 to 90·4) | 33·8<br>(22·8 to 46·3) |
| **sTREM-1 model** | | | | | | |
| 0·1 | 87·2<br>(72·6 to 95·7) | 29·1<br>(21·4 to 37·9) | 0·44<br>(0·19 to 1·04) | 1·23<br>(1·04 to 1·45) | 88·1<br>(74·4 to 96·0) | 27·4<br>(19·8 to 36·2) |
| 0·15 | 74·4<br>(57·9 to 86·9) | 46·5<br>(37·6 to 55·5) | 0·55<br>(0·31 to 0·97) | 1·36<br>(1·09 to 1·77) | 85·5<br>(74·9 to 92·8) | 29·9<br>(21·0 to 40·0) |
| 0·20 | 61·5<br>(44·6 to 76·6) | 63·8<br>(54·8 to 72·1) | 0·60<br>(0·40 to 0·92) | 1·7<br>(1·21 to 2·38) | 84·4<br>(75·5 to 90·9) | 34·3<br>(23·4 to 46·6) |

**Supplementary Figure 4. Sensitivity and specificity for each model in the validation cohort**. Coloured lines indicate point estimates and pink shaded areas indicate 95% confidence intervals.

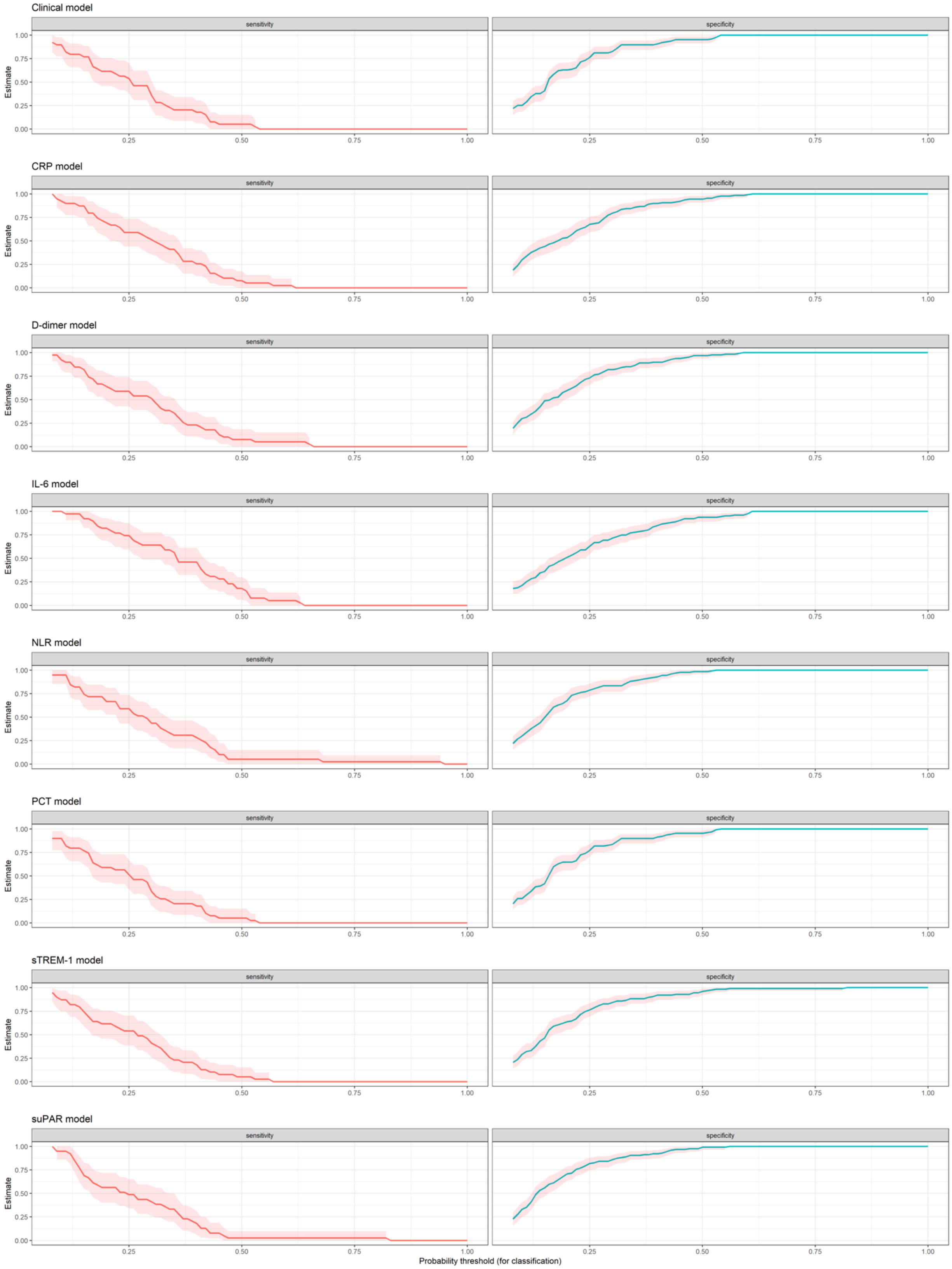

**Supplementary Table 7. Association of corticosteroid use with primary outcome.** Corticosteroid use is defined as use of oral or parenteral steroids occurring at least one calendar day prior to developing an oxygen requirement for participants who met the primary outcome.

| | | Developed oxygen requirement | | | Relative risk (95% CI) | p-value |
|---|---|---|---|---|---|---|
| | | Yes | No | TOTAL | | |
| **All participants** | | | | | | |
| **Corticosteroid use** | Yes | 31 | 127 | **158** | 0·90 (0·61-1·32) | 0·58 |
| | No | 58 | 207 | **265** | | |
| | **TOTAL** | **89** | **334** | **423** | | |
| **AIIMS participants** | | | | | | |
| **Corticosteroid use** | Yes | 12 | 53 | **65** | 0·73 (0·37-1·42) | 0·35 |
| | No | 15 | 44 | **59** | | |
| | **TOTAL** | **27** | **97** | **124** | | |
| **CMC participants** | | | | | | |
| **Corticosteroid use** | Yes | 19 | 74 | **93** | 0·98 (0·60-1·58) | 0·93 |
| | No | 43 | 163 | **206** | | |
| | **TOTAL** | **62** | **237** | **299** | | |

**Supplementary Table 8. Candidate predictor variables, stratified by corticosteroid use.** *Missing data: CRP = 8, D-dimer = 3, IL-6 = 2, NLR = 12; PCT = 2; sTREM-1 = 2. Median values (IQR) and p-values for Wilcoxon rank sum tests are reported for continuous variables. Pearson's Chi-squared test p-values are reported for categorical variables.

| **Candidate predictor** | **Overall** (n = 423) | **Corticosteroid use** | | **p-value** |
|---|---|---|---|---|
| | | **No** (n = 265) | **Yes** (n = 158) | |
| Age (years) | 53·0 (41·0 to 62·0) | 53·0 (40·0 to 62·0) | 54·5 (42·0 to 62·0) | 0·4 |
| Male sex | 286 / 423 (68%) | 175 / 265 (66%) | 111 / 158 (70%) | 0·4 |
| Oxygen saturation (%) | 98·0 (96·0 to 99·0) | 98·0 (96·0 to 99·0) | 97·0 (96·0 to 98·0) | 0·017 |
| CRP (mg/l) * | 36·7 (7·0 to 108·6) | 37·4 (6·8 to 110·3) | 35·5 (7·3 to 101·0) | 0·8 |
| D-dimer (ng/ml) * | 856·1 (470·8 to 1,522·7) | 846·7 (484·0 to 1,484·5) | 882·1 (458·4 to 1,706·4) | 0·6 |
| IL-6 (pg/ml) * | 19·8 (6·6 to 47·6) | 21·2 (7·2 to 53·9) | 15·4 (4·8 to 43·2) | 0·042 |
| NLR * | 3·1 (1·9 to 4·9) | 2·7 (1·7 to 4·3) | 3·7 (2·3 to 6·2) | < 0·001 |
| PCT (ng/ml) * | 0·1 (0·1 to 0·2) | 0·1 (0·1 to 0·2) | 0·1 (0·1 to 0·2) | 0·8 |
| sTREM-1 (pg/ml) * | 394·0 (272·0 to 563·0) | 400·0 (269·0 to 563·0) | 381·0 (281·2 to 565·2) | > 0·9 |
| suPAR (ng/ml) | 4·2 (3·1 to 5·7) | 4·1 (3·0 to 5·6) | 4·3 (3·2 to 5·8) | 0·2 |

**Supplementary Table 9. Association of baseline Ct value with primary outcome, stratified by collection technique, PCR platform or both**. Nasopharyngeal swabs (NPS) were collected at CMC and combined nasopharyngeal and oropharyngeal swabs (OPS) were collected at AIIMS. Only swabs collected within the 24 hours prior to recruitment (n = 242/423) are included in this sub-analysis. All swabs were tested using the Cepheid Xpert Xpress SARS-CoV-2, CA at AIIMS. Both the Cepheid Xpert Xpress SARS-CoV-2, CA and Altona RealStar SARS-CoV-2 rRT-PCR, Hamburg were used at CMC. Median values (IQR) and p-values for Wilcoxon rank sum tests are reported.

| Stratification factor | Ct value | | | p-value |
|---|---|---|---|---|
| | Overall | Developed oxygen requirement | | |
| | | No | Yes | |
| All participants (n = 242) | 27·7 (23·1 to 32·7) | 27·4 (22·8 to 33·0) | 28·4 (24·1 to 32·4) | 0·4 |
| **Collection technique (n = 242)** | | | | |
| NPS (n = 118) | 31·5 (26·7 to 35·7) | 31·5 (26·1 to 35·8) | 31·5 (28·0 to 34·6) | 0·9 |
| Combined NPS and OPS (n = 124) | 25·3 (20·5 to 28·5) | 25·5 (20·2 to 28·8) | 24·5 (21·4 to 27·8) | > 0·9 |
| **PCR platform (n = 242)** | | | | |
| Cepheid Xpert Xpress (n = 173) | 26·9 (21·0 to 31·5) | 26·6 (20·7 to 30·9) | 27·6 (24·0 to 32·1) | 0·2 |
| Altona RealStar (n = 69) | 31·6 (27·0 to 35·4) | 31·8 (27·0 to 35·4) | 31·2 (28·0 to 35·4) | 0·6 |
| **PCR platform at CMC site (n = 118)** | | | | |
| Cepheid Xpert Xpress (n = 49) | 31·2 (26·3 to 35·8) | 30·7 (23·2 to 36·1) | 31·9 (28·4 to 33·3) | 0·4 |
| Altona RealStar (n = 69) | 31·6 (27·0 to 35·4) | 31·8 (27·0 to 35·4) | 31·2 (28·0 to 35·4) | 0·6 |
